# Reflective Learning from Implementing Care Pathways for Vulnerable Infants and Their Mothers: Case studies from Pakistan, South Sudan and Yemen

**DOI:** 10.1101/2024.12.02.24318218

**Authors:** Hedwig Deconinck, Stephanie V. Wrottesley, Saba Shahid, Angelina Nasira Boi, Pascal Mane Luka, Marlene Traore Hebie, Kirk Dearden, Ahmed Aljabi, Emily Hirata, Marie McGrath

## Abstract

Many infants are born vulnerable or become vulnerable in the first 6 months of life, putting them at risk of poor growth and development, long-term ill-health, and increased mortality. Many policies and practitioners aim to integrate care across services for at-risk mother infant pairs, but this is challenging in practice given individual, setting, and policy complexities. The MAMI Care Pathway, an integrated care pathway approach, frames and guides implementation of WHO nutrition and health guidelines within health and nutrition systems. Local contextualization is a pre-requisite for evidencing national policy uptake of relevant global recommendations. To gain insight into such contextualization, we undertook three in-depth country case studies to investigate planning, implementation, adoption, and scalability of the approach with practitioners in Pakistan, South Sudan, and Yemen.

Our mixed methods using theoretical frameworks explored planning, implementation, normalization, and sustainable scalability of the care pathway. We explored healthcare workers’ perspectives through in-and cross-country reflective learning discussion.

Initiatives were prompted by perceived but unmeasured need. Planning and starting implementation required substantial time and stakeholder consultation. Healthcare workers needed skills to perform existing tasks (reflecting shortfalls in routine care capacity and quality) as well as new ones (e.g. maternal mental health). Shifting from disease-focused to person-centred care required (and did not always get) buy-in from healthcare workers (where it was not embedded in service duties and pay), and from communities and mothers to adopt it (practical and competing priorities). It was possible to be system-supportive and scale-sensitive in emergency contexts.

Identified commonalities in implementation realities heighten transferable learning potential. Health system strengthening is needed for impactful scale. A paradigm shift towards person-centred care is needed in policy framing and implementation guidance. Deeper localized case study development is a critical type of evidence to inform policy navigation, development and practice.

**TEASER KEY MESSAGE:** Reflective learning from adapting, implementing, and normalizing the MAMI Care Pathway in Pakistan, South Sudan, and Yemen health systems highlighted the need for system-supportive, scale-sensitive approaches and financial and technical support to align with national policies and implementation realities.

**KEY MESSAGES:**

- Infants born vulnerable or becoming vulnerable early in life risk poor growth and development, ill-health, and mortality. Integrated care for at-risk mother-infant pairs is the goal of many policies but is challenging in practice. Case studies in Pakistan, South Sudan, and Yemen investigated the process of adapting, implementing, and normalizing the integrated MAMI Care Pathway approach by putting WHO guidelines on outpatient care of at-risk infants and their mothers into practice to inform feasible, sustainable care.
- The cases in these diverse contexts demonstrated that system strengthening, and early healthcare provider engagement were crucial for impactful and sustainable scale of the MAMI Care Pathway approach. Identifying commonalities in implementation realities enhance transferable learning.
- Embedding a sustainable approach in existing health services requires financial and technical support aligning with national health policies, planners, and implementation. Achieving comprehensive care also demands investment in routine service capabilities.
- A paradigm shift toward person-centred care is needed in policy and implementation guidance. Deeper localized case studies are critical evidence to inform policy navigation, development, and practice.

## BACKGROUND

### Infant vulnerability

Many infants are born vulnerable or become vulnerable in the first 6 months of life, putting them at risk of poor growth and development, long-term ill-health, and increased mortality.^1^ Annually, 8.9 million at-risk babies (15.0%) are born with low birth weight (LBW), which carries short-and long-term risks, especially for premature babies.^2^ In low- and middle-income countries (LMICs), 9.2 million (15.5%) infants under 6 months of age (u6m) are wasted, 10.3 million (17.4%) are underweight, and 11.8 million (19.9%) are stunted.^3^ Early life wasting increases the risk of malnutrition, contributing to the global burden of 45 million children under 5 years of age who are wasted and 149 million who are stunted,^1^ affecting health outcomes in current and future generations.^4, 5^

Vulnerable infants u6m may be described as newborns with LBW due to preterm or small for gestational age; infants nutritionally at-risk or identified with wasting, stunting, or underweight; infants with acute or chronic illness, disability, or other growth and development concerns; and infants whose mothers have nutrition, physical, mental health, or social challenges.^6^ Comprehensive, personalized care for these infants and their mothers is essential but challenging because of healthcare system complexity.^7, 8^

### Global policy context

Multiple global guidelines describe what to do to care for vulnerable infants u6m.^6^ The “how” includes navigating policy recommendations and developing implementation guidance contextualized to settings, including the policy environment.

In 2015, the MAMI Care Pathway^9, 10^ was developed as an adaptable framework and resource package to apply the 2013 WHO recommendations on outpatient care of malnourished infants u6m^11^ and generate implementation evidence to inform policy guidance development. Implementation learning has informed the expansion of the 2023 WHO guideline on child wasting^12^ to include the management of infants u6m at risk of poor growth and development, as well as their mothers, embedding core components of the MAMI Care Pathway approach.

WHO has identified the MAMI Care Pathway as a “candidate intervention” in implementation guidance development for small and sick newborn follow-up, feeding of at-risk infants u6m, and risk-differentiated personalized care. The approach strongly aligns with current WHO policy development and updates, including the integrated management of childhood illness (IMCI).^13^

### National policy uptake

Despite progress at the global policy level, governments have been slow to apply the 2013 WHO recommendations because of lack of contextualized evidence and limited resources. The shift from inpatient care (pre-2013) to outpatient care (2013), along with the expansion of the target group (2023) has considerable implications for service capacity but can help realize integrated care within healthcare systems.

Feasible, sustainable care is a pre-requisite for successful and responsible scale-up. To ensure an intervention’s effectiveness and scalability, it is crucial to gather evidence not only on its impact but also on the conditions under which it is implemented to ensure system “fit” and inform pathways to scale.^14-16^ This approach enhances replicability and ensures sustainable delivery. Learning from small-scale implementation before expanding is crucial, necessitating meticulous planning from the outset.^17^

### Implementation evidence

Through the MAMI Global Network^18^ the MAMI Care Pathway approach has continuously evolved. Given its alignment with global policy recommendations and the need for implementation evidence from national stakeholders, ENN, as network coordinator, developed a series of in-depth “learning by doing, together” case studies in collaboration with national implementers.

Our case study series generated learning from implementation of the Care Pathway approach in three contexts to inform approaches to sustainable scalability of care. The series asked (a) what was done, how, and why, (b) what practices worked and did not work, for whom and under what circumstances, (c) how the approach was spread, scaled up and sustained, and (d) how practices improved and were sustained at scale.

## METHODS

We used mixed methods for iterative and participatory reflective learning and applied different lenses to explore experiences within and across settings to inform transferability.^19^ The realist helped deal with context and complexity.^19, 20^ Four frameworks guided the inquiry. We explored embedding care by applying the *Normalization Process Theory (NPT)*.^21, 22^ The spread, scalability, and sustainability of care inquiry used the *Non-adoption, Abandonment, Scale-up, Spread and Sustainability (NASSS) Framework*^23^ and the *Checklist for Assessing the Potential Scalability of Pilot Projects or Research*.^17 24^ We developed a Planning and Implementation *Process Framework specific for the MAMI Care Pathway Approach* for each context (*Supplemental Materials 1, Methods*).

### Selection of sites and respondents

We looked for different implementation modalities in different regions, in rural and urban settings of LMICs, considering contexts, implementers, geography, data availability, and transferable learning potential. We selected a pediatrician-led clinic in a tertiary hospital in Pakistan, an implementation study in urban and rural maternal and child health (MCH) services in South Sudan, and an implementation pilot in a health and nutrition emergency program in Yemen. Respondents (co-learners and co-authors) were sub-national health and nutrition managers and practitioners from ministries of health (MOH), academic, private and non-governmental organizations (NGOs). All respondents provided informed consent.

### Data collection and tools

We developed generic interview tools for the inquiry and adapted them to each country case. For the “planning and implementation” investigation, we asked participants to respond to email questionnaires. Oral feedback and clarification sessions focused on the planning process and social dimensions. We then explored with how to normalize the MAMI care pathway in their routine practitioners’ work. Next, we discussed the sustainability of the care pathway with senior managers and clinical healthcare workers across contexts and settings. These discussions allowed us to triangulate learning and reflect on potential scalability, detect commonalities, and generate transferable insights. During data collection, we recorded biases, interference, or limitations. Data tools included 1) a questionnaire on planning and implementation, 2) an interview guide on normalizing the care pathway in routine services, 3) a checklist on non-adoption, abandonment, scale-up, spread, and sustainability for group discussions, and 4) a checklist to assess potential scalability (*Supplemental Materials 2, Data Tools*).

### Data analysis

We used an iterative approach to evaluate case experience, applying repeated cycles of testing and theory building. Methods were both deductive (testing ideas) and inductive (discovering new ideas). Three kinds of analysis generated practical, pragmatic guidance to improve care. “Theory-driven” analysis examined planning, introduction, adaptation, implementation, monitoring, and improvement of the approach and assessed readiness for scale-up; explorative data analysis investigated clinical healthcare workers’ perceptions, identified what worked for whom and under what circumstances, and evaluated adoption; and explanatory analysis of all available data allowed us to triangulate findings and deepen our understanding.

### Participatory and adaptive, reflexive learning

Participants engaged in reflective “learning together by doing” that deepened their understanding of how to embed and adapt the MAMI Care Pathway approach in local health systems. This method tapped tacit knowledge and contributed significantly to collective learning.

### Patient and public involvement

No patients or public were involved.

## RESULTS

First, we described the health context in each country. Second, we described planning and implementing care—what was done, how, and why—and measured outcomes. Third, we explored factors that influenced individual and collective actions to adopt (normalize) the approach in routine services. Finally, we evaluated the potential scalability and sustainability of the approach.

### 1. Prevailing health contexts by setting

Despite different settings (one non-emergency, two protracted emergencies), the three countries had similar health indicators (Box 1). Data on the burden of vulnerability of infants u6m were not available, but based on experience, we expected large numbers of infants at risk of or experiencing poor growth and development, with mothers needing additional care and support. National policies recommended referring infants u6m with severe wasting or nutritional oedema to hospital for inpatient care, with no outpatient care option. Local health systems in Yemen and South Sudan, typically supported by international NGOs, had limited and interrupted funding, making service delivery unstable. A notable difference in skilled birth attendance was highest in Pakistan (74%) (2019),^25^ lowest in South Sudan (19%) (2010),^26^ and in between in Yemen (45%) (2023).^27^ Regarding burden, the population of Pakistan is 17 times that of South Sudan and 7 times that of Yemen.

#### Box 1.

**Health context by setting**

In **Pakistan**, maternal mortality had halved since 1990,^28^ but substantial disparities in health access still affect vulnerable women and children the most. Neonatal and infant mortality rates were high, 41 and 55 deaths per 1,000 live births respectively, with 19% of infants born with LBW and 49% of infants exclusively breastfed.^25 29^ In children 6-59 months, wasting and stunting rates were 18% and 40%, respectively (2018).^29^ Data were unavailable for infants u6m.

In **South Sudan**, 96% of the population live in rural areas, and 56% lack access to healthcare services. Among people with access to healthcare services, 54% pay out of pocket. In 2021, an estimated 8 out of 14 million people needed humanitarian assistance.^30^ Rates of neonatal and infant mortality were 40 and 64 deaths per 1,000 live births, respectively (2021).^31^ Historic national data showed that 74% of infants u6m were exclusively breastfed (2018). ^32^ In children 6-59 months, 13% were wasted and 4% severely wasted over the past decade, and 17% were stunted (2019).^33^ The healthcare system had strong national Ministry of Health (MOH) leadership but inadequate resources (e.g., skilled healthcare professionals), and international non-governmental organizations (NGOs) and donors ran or supported implementation of services.

In **Yemen**, the protracted crises of 2023 put an estimated 24 of 34 million people at risk of hunger and disease, and 14 million in need of humanitarian assistance. Neonatal and infant mortality rates were 28 and 47 deaths per 1,000 live births, respectively (2023).^27^ Only 10% of infants were exclusively breastfed (2023).^27^ In children 6-59 months, wasting and stunting rates were 18% and 34%, respectively (2019).^27^ International NGOs and donors supported the MOH in fragile healthcare system areas.

### 2. Planning and implementing the MAMI Care Pathway to what effect by setting

The modes of initiating and determining implementation varied across different contexts (Box 2). During a collaborative planning and implementation process, the MAMI care pathway activities were discussed, and decisions were made regarding what was done where, how, and by whom, based on the existing policies and capacities in each context. *Supplemental Materials 3 MAMI Care Pathway Unpacked* maps the care pathway activities across the three contexts, showing minimal variation. Respondents agreed that this mapping was crucial to understand capacity gaps and needs, avoid duplication, and optimize operations and resources. It helped healthcare workers make the right decisions, assume ownership and responsibility, and visualize comprehensive care with continuity between services and over time in their settings.

#### Box 2.

**Implementation prompt and modality by setting**

In **Pakistan**, the pediatric outpatient department (OPD) of the Indus Hospital in Karachi, a charitable, free tertiary teaching hospital, introduced the MAMI care pathway approach in 2021 in response to an identified gap in care. The chair of pediatrics spearheaded the introduction, receiving approval from the hospital medical directorate to develop a MAMI policy, establish a new unit, and provide the needed resources and support. She sought support from MAMI Global Network experts to contextualize materials and trained clinical healthcare workers and support staff. It took four months to receive approval and set up the clinic. Enough healthcare workers were allocated to run the clinic three times a week, receiving a maximum of 18 vulnerable mother-infant pairs as outpatients, referred from pediatric and maternity services in the hospital. Pediatric residents rotated through the clinic as part of their training and were mentored and supported by the pediatric supervisor.

In **South Sudan**, the MAMI care pathway approach was introduced in 2021 through a 15-month implementation study, as part of a multi-year non-governmental organization (NGO)-led maternal and child health (MCH) project. The MOH and the donor agreed to make existing resources available as part of ongoing MCH services support. The MOH purposively selected one site in each of four districts in four states to represent the national healthcare system in diverse rural and urban settings. Each site consisted of an MOH-run primary healthcare centre (PHC), catchment communities, a referral hospital, and the County Health Department (CHD) for oversight. Additional NGO staff and resources were allocated for implementation support, training, and monitoring and evaluation (M&E). An integrated approach was planned from the start, building on existing healthcare services as much as possible in an implementation research context. An initial situation analysis included policy, stakeholder, and local health system capacity assessments. In participatory discussions and through implementation, national decision makers, managers, and trained healthcare workers adapted, tested, and refined generic MAMI care pathway materials. It took nine months for the NGO to access funding, plan, gain MOH ethical committee approval, and set up the sites. An external consultant supported the planning and implementation.

In **Yemen**, the MAMI care pathway approach was introduced in 2021 through a multi-year NGO-led health and nutrition emergency program in nine MOH-run PHCs in four districts across three governorates, addressing a perceived need in a highly vulnerable population. Additional NGO staff were allocated to support implementation, and existing resources were made available as part of ongoing MCH and nutrition services support, in agreement with the MOH and the donor. The established relationship and existing expertise facilitated start-up within six months. An external consultant briefly supported the planning.

Each country used similar criteria to screen, assess, and classify the risk of vulnerable infants u6m and their mothers (*Supplemental Materials 4 MAMI Care Pathway Criteria)*. Similar quantitative data were gathered across the three settings. Table 1 compares the key indicators for screening, assessment, enrolment, and outcomes of vulnerable mother-infant pairs.Pakistan had the highest proportion of pairs assessed with moderate risk (65%), South Sudan the lowest (27%), and Yemen in between (48%). Among all the pairs assessed, 5% were found to be at high risk in South Sudan and 3% in Yemen. In terms of moderate risk, 65% were found in Pakistan, 27% in South Sudan, and 48% in Yemen. The low proportion of pairs classified as high risk at the PHC may mask the true situation in the community. In South Sudan, only 32% of pairs were at moderate or high risk. This low percentage may suggest that the screening method was not specific enough, resulting in an unnecessarily high burden for assessments. This may also explain why fewer pairs were enrolled out of those assessed in South Sudan than in Pakistan and Yemen. Enrollment had slightly favored boys in Pakistan and girls in Yemen. Outcomes across sites were relatively similar despite variation in exit criteria. Recovery (proportion of enrolled pairs who had retained in care until the infants reached 6 months of age with resolved risks) was better in South Sudan (84%) and Yemen (70%) than in Pakistan (60%). Missingness (proportion of enrolled pairs who missed to retain in the care pathway until the infant reached 6 months of age due to death or loss to follow up), was very high in South Sudan (58%) and lower in Yemen and best in Pakistan (36% and 26% respectively). Contributing factors may include weak community engagement, recruitment outside the health catchment area, constraints to maternal attendance, and high population mobility due to insecurity, work, or seasonal moves.

**Table 1.** Screening, assessment, enrolment, and outcomes of mother–infant pairs for a 12-month period, Pakistan, South Sudan and Yemen cases, 2023.

|  | **Pakistan case** | **South Sudan case** | **Yemen case** |
| --- | --- | --- | --- |
| **Period covered:** | December 2021–2022 | October 2022–September 2023 | July 2022–June 2023 |
| **Key indicators:** |  |  |  |
| **Pairs screened** | NA | 4,813 | 4,615 <sup>a</sup> |
| **Pairs screened identified at risk** | NA | 1,971 | 2,749 <sup>a</sup> |
| **Pairs assessed** | 1,238 | 1,971 | 1,866 |
| **Pairs assessed identified at moderate risk (% of pairs assessed)** | 807 (65.2%) | 529 (26.8%) | 891 (47.7%) |
| **Pairs assessed identified at high risk (% of pairs assessed)** | NA | 94 (4.7%) | 60 (3.2%) |
| **Pairs assessed boy/girl ratio** | 1.23 | 1.03 | 0.84 |
| **Key reasons infants' moderate risk** | Mixed feeding, slow weight gain | LBW, low MUAC, feeding difficulties | LBW, slow weight gain |
| **Key reasons mothers' moderate risk** | Work-related absence | Adolescent motherhood | Adolescent motherhood |
| **Pairs enrolled in care** | 807 | 521 | 891 |
| **Pairs recovered at infant aged 6 months (% pairs attending care until infant aged 6 months)** | MUAC: 358 (59.6%)<br>WAZ: 447 (74.4%) | 183 (83.6%) | 402 (70.3%) |
| **Pairs not recovered at infant aged 6 months (% of pairs attending care until infant aged 6 months)** | MUAC: 243 (40.4%)<br>WAZ: 154 (23.6%) | 36 (16.4%) | 170 (29.7%) |
| **Pairs missed before or at infant aged 6 months (died, absented, did not return, lost to follow-up) (% of pairs enrolled)** | 206 (25.5%) | 302 (58.0%) | 319 (35.8%) |

<sup>a</sup> Data represent a 10-month period, October 2022–July 2023.
LBW= low birth weight; MUAC= mid-upper arm circumference; NA= not available; WAZ= weight-for age z score.

Monitoring, learning, and reporting for quality improvement varied widely across the country cases. In Pakistan, a digital MAMI monitoring system was added to the hospital’s fully digitized medical record system. This generated post-hoc implementation learning but was not accessible to healthcare providers for real-time feedback on quality of care or professional development. In South Sudan, for learning and quality improvement, a partially digitized spreadsheet with automated analyses monitored monthly progress and quality of care with implementers. In Yemen, activities and progress were mainly captured for donor reporting.

### 3. Embedding care in routine services

We assessed the adoption of the MAMI care pathway in routine practice through coherence, cognitive participation, collective action, and reflective monitoring, identifying promoting and hindering factors.

In **Pakistan**, making rotation in the MAMI clinic mandatory as part of the pediatric residency program ensured strong support from supervisors, which motivated residents to excel. Assessing, monitoring, and addressing infant and maternal risk factors that affected infant growth and development over time required clinicians to develop new skills to guide and motivate mothers to adhere to care protocols.

In **South Sudan**, clinicians were supported to understand the care approach, acquire necessary skills, and engage in quality improvement and continuous learning. However, they perceived the new approach as an additional task and expected remuneration. They were unlikely to adopt it in their daily work unless it was included in their job description and career development plans. Shifting from disease-focused to person-centred care was challenging in the country’s vertical, program-driven health system. Staff did not value benefits such as reduced workload through streamlined management, better resource management, or improved teamwork and task distribution.

In **Yemen**, clinicians received a small remuneration as motivation to take on additional workload. They were gratified to offer care to a population that was previously ignored or sent to hospital. Although shifting from disease-focused to person-centred care of mother-infant pairs did not reduce their workload or improve teamwork, they appreciated their increased expertise.

We evaluated the likelihood of clinicians making care routine by plotting the adoption scores across the adoption components. Results were generally positive in all three cases (figure 1). In Pakistan, reflective monitoring scored low, but discussions indicated that an improved monitoring system was being developed. In South Sudan, cognitive participation was low because clinicians felt they were testing a new intervention rather than adopting a national recommendation, and collective action scored low because few clinicians were involved in care per site. In Yemen, reflective monitoring scored low due to the lack of an established monitoring system, and cognitive participation and collective action were rated low because few clinicians were involved and implementing the care pathway was seen as a job requirement.

**Figure 1:**
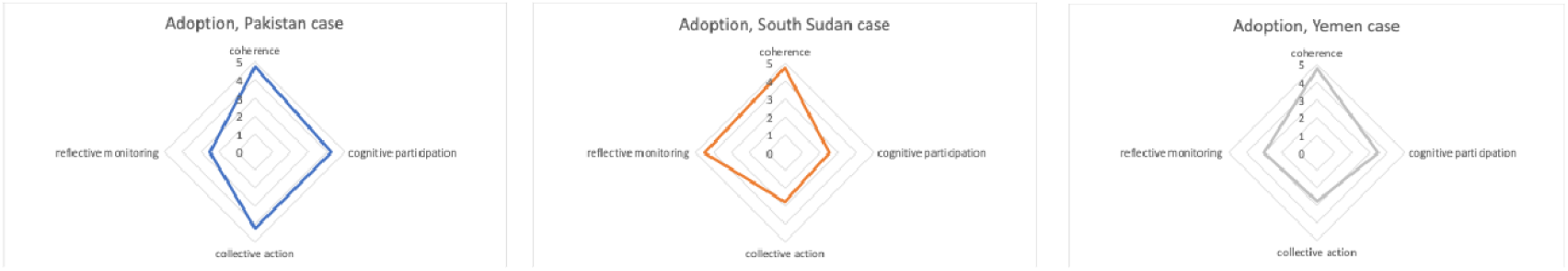
Interpreting the adoption of the MAMI Care Pathway approach, Pakistan, South Sudan and Yemen cases, 2023. Adoption was scored on a sliding scale from 1 “not adopted at all” to 5 “completely adopted”.

### 4. Spread, scalability, and sustainability of care

Our first method identified **challenges and factors predicting the success of scaling the MAMI care pathway**. Reflective discussions across seven domains (condition, technology, value proposition, adopters, health organization, wider system, and adaption over time) generated insights for scalability. We graded these domains as (1) simple or easy, (2) complicated, more difficult, and (3) complex, challenging. The results, presented as radar figures, showed variation across the country cases (figure 2). Overall (summed) feasibility scores were similar (Pakistan 16, South Sudan 18, Yemen 15), though the content varied.

**Figure 2:**
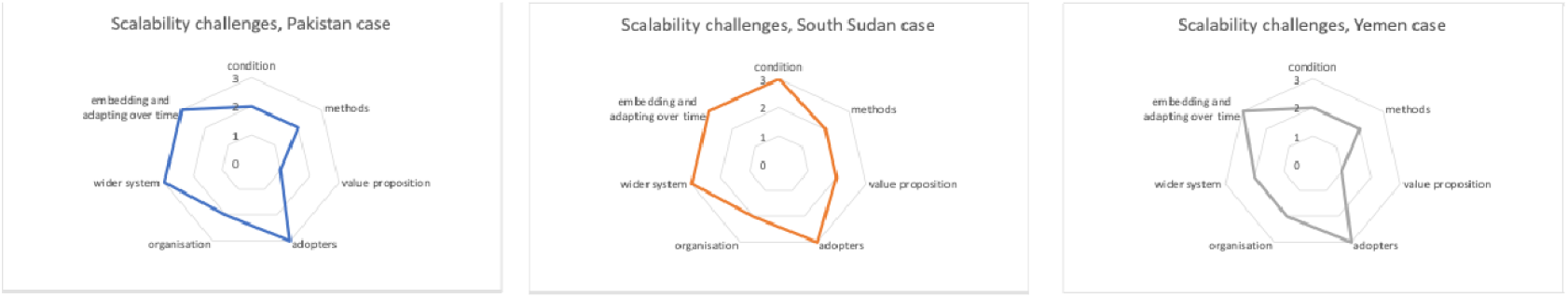
Appraising challenges to the scalability of the MAMI Care Pathway implementation, Pakistan, South Sudan and Yemen cases, 2023. Challenges were graded as 1 “simple”, 2 “complicated”, and 3 “complex” to address.

In **Pakistan**, pediatric residents understood the descriptor “small and nutritionally at-risk infants and their mothers”, though some risks outside their comfort zone (e.g., maternal mental health, socioeconomic factors) were harder to detect, and the mother–infant pair focus was new (grade 2). The methods for assessing and providing care was mostly known, but integrating maternal health required new skills (grade 2). Both residents and mothers understood the benefit (value proposition) (grade 1). However, clinicians, adopters-to-be, initially resisted taking on additional tasks, and mothers, who valued the service, faced barriers to return for follow-up visits (grade 3). Strong leadership and resources were needed to establish the clinic (grade 2). The wider system faced challenges in expanding and regularizing the approach because of competing health priorities (grade 3). Without policy changes, embedding and adapting the approach over time faced significant barriers (grade 3).

In **South Sudan**, despite skills training, intense coaching and job aids, healthcare workers found some risk factors (e.g., disabilities, maternal mental health) difficult to detect. The mother-infant pair focus was new and influenced by socio-cultural factors and co-morbidities (grade 3). They were familiar with methods to assess and support at-risk infants, but focusing on mother-infant pairs required new skills and contextual adaptation (grade 2). Most but not all healthcare workers valued the benefit of identifying vulnerability early (grade 2). Introducing the care pathway changed staff roles and increased workloads, while mothers’ acceptance depended on perceived benefits and support networks (grade 3). Implementing the care pathway required significant organizational changes and leadership, which was challenging in a resource-limited context (grade 2). While national interest in the MAMI care pathway grew, more financial and policy support was needed to align it with existing programs and support the paradigm shift to person-centred care (grade 3). Continuous learning and alignment with other approaches over time were needed to overcome significant barriers (grade 3).

In **Yemen**, healthcare workers understood the condition adapted to primary care, but risks of disabilities and maternal mental health were hard to detect (grade 2). Methods to assess and support at-risk infants with a mother-infant pair focus, adding maternal factors, required new skills and adaptations (grade 2). Healthcare workers valued the early detection and support for at-risk mother-infant pairs, and consistent communication strengthened mothers’ understanding and confidence (grade 1). Staff roles changed with increasing workloads, and mothers found the process burdensome, but both gained confidence (grade 3). Integrating care into existing services required significant organizational changes and external support (grade 2). While national interest grew, financial, technical, and policy support were needed to align with existing programs and manage competing health priorities (grade 3). Embedding and adapting care over time require a more robust monitoring system to support quality improvement and national learning (grade 3).

Our second method explored **potential scalability by assessing readiness for scale-up**. We considered key actions in the design that enhance potential large-scale impact ^17^. We explored and mapped twelve key actions across the country cases (table 2), indicating whether these actions were taken or missed, with confirmation (yes) indicating progress or improvement rather than full completion.

**Table 2:** Appraising potential scalability of the MAMI Care Pathway implementation, Pakistan, South Sudan and Yemen cases, 2023.

| Appraisal of actions for sustainable scale-up | Pakistan case | South Sudan case | Yemen case |
| --- | --- | --- | --- |
| 1. Involved future stakeholders | No | Yes | Yes |
| 2. Addressed a persistent health condition or service | Yes | Yes | Yes |
| 3. Considered expectations about scale-up in the design | No | Yes | Yes |
| 4. Considered constraining or supporting socio-cultural and gender factors | No | Yes | Yes |
| 5. Kept package of interventions simple, without jeopardizing outcomes | No | Yes | No |
| 6. Tested in a variety of socio-cultural and geographic settings | No | Yes | No |
| 7. Required no extra human and financial resources for implementation | No | No | No |
| 8. Assessed and documented health outcomes and process of implementation | Yes | Yes | Yes |
| 9. Engaged with donors and technical partners to support scale-up early and continuously | No | No | No |
| 10. Planned to advocate for changes in policies and regulations | No | Yes | No |
| 11. Designed mechanisms to review progress and incorporate new learning | Yes | Yes | Yes |
| 12. Shared understanding on the importance of adequate evidence on feasibility and outcomes prior to scale-up | Yes | Yes | Yes |

In **Pakistan**, only four out of twelve actions were achieved, because the implementation mode was initially designed to make services available routinely and sustainably in a single hospital setting, without scale-up in mind. Consequently, recommended actions for scaling up were not applicable. However, desire was expressed to extend the MAMI care pathway approach to satellite clinics, which would require policy changes and additional commitment and resources. Healthcare workers were interested in sharing the learning with similar hospital settings and planned peer-reviewed publications to support this effort and attract the attention of pediatric colleagues.

In **South Sudan**, all but two key actions were considered. *Not requiring extra human and financial resources* (action 7) was not achieved because the study was meant to test and learn from implementation under existing resource constraints. While the local health system theoretically had the capacity to implement the care protocol, external support was brought in for testing and learning. *Engaging with donors and technical partners to support scale-up early and continuously* (action 9) was planned but had not started, despite regular donor updates and support for implementation by other NGOs in the country. Initially, in-country health and nutrition actors showed little interest in collaborating and learning about the implementation, despite their agency’s participation in the MAMI Global Network. However, this situation began to change with increasing visibility of the approach and support from donors.

In **Yemen**, five actions were not achieved. *Keeping the package of interventions as simple as possible, without jeopardizing outcomes* (action 5) was not achieved because generic care materials received only minor changes to align with existing care because the emergency context favored rapid implementation and in-house capacity was limited. *Testing the innovation in a variety of sociocultural and geographic settings* (action 6): The emergency project enabled testing of the innovation in the NGO-supported sites without adapting to each specific context. *Not needing extra human and financial resources for implementation* (action 7): The project allowed additional staff and salary top-ups to compensate for increased tasks and workload. *Seeking early and continuous financial support from donors and technical partners for scale-up* (action 9): All activities relied on emergency funding, which could change or end with shifting donor priorities, but no sustainable development funding was sought. *Planning for advocacy for changes in policies and regulations* (action 10): No plans were developed, although the NGO recognized the need and expressed a desire to engage with the state MOH. *Assessing and documenting the implementation process and health outcomes (*action 8), and *Instituting mechanisms to review progress and incorporate new learning into the implementation process* (action 11) were weak, indicating deficiencies in the monitoring and learning system. Nevertheless, the NGO successfully used time-limited emergency funding to gain initial experience and continue (with repeated short-term grants) and offered their expertise to other health actors in the country.

## DISCUSSION

The case studies generated rich “real world” insights into the needs and potential for planning, implementing, adopting, and scaling up the MAMI Care Pathway approach across the three countries.

**The Pakistan case** demonstrated the feasibility of integrating the care approach in a tertiary teaching hospital with good leadership, flexible hospital management, and support for resident pediatricians to adapt practices. Lack of access to satellite primary care and community health systems closer to the vulnerable households constrained sustainable support. Also, reaching colleagues and influencing major health actors, including the State MOH and WHO, was difficult for hospital healthcare providers. **The South Sudan case** demonstrated the feasibility of implementing the integrated care approach building on existing maternal and child health services in collaboration with the MOH. Additional resources were required for the implementation research to adhere to the study protocol and produce robust results, and this limited the immediate feasibility of sustainable integration into the existing local health system. Current national policies and practices would require revision to accommodate this. The start-up was slow, but preparations were thorough. The strategy and learning provided valuable insights to inform discussions on policy adjustments necessary for future integration and scale-up. **The Yemen case** demonstrated the feasibility of swiftly integrating the care approach into an emergency health and nutrition emergency intervention using flexible, piggy-backed short-term (humanitarian) resources. Partners, including the MOH, highly valued the learning because it addressed a need. This sparked interest in adopting the care approach among other emergency health and nutrition projects, for which the NGO helped build capacities. Strengthened monitoring and documentation would have facilitated this process.

Across all three countries, the efficiency of MAMI screening methods could be further investigated to optimize for false positives and false negatives to minimize workload from lengthy assessments. The excessive number of cases lost to follow-up during the intended treatment period (infants reaching 6 months of age) requires immediate attention.

**Key insights** generated during case discussions from different country perspectives were remarkably similar. This sparked transfer of learning between country co-learners during the case studies and heightens transferability potential in and beyond these settings.

Insights from healthcare workers based on their experience with vulnerable mothers:

- Mothers are motivated by seeing their babies’ healthy growth, which boosts their self-confidence and self-respect (“we can do it”) and increases their trust in healthcare workers and the health system.
- A broader support system improves community awareness and engagement to support vulnerable mother-infant pairs, informing and enabling them to change risky behaviours and address barriers to accessing care.

Insights from service implementers:

- Local healthcare teams of multiple experts working together with a vision of integrated, person-centred continuity of care foster responsibility, motivate staff, and alleviate workload. This requires adapting care procedures and linking with specialized services for referring cases within and outside the health facility.
- Collaboration with health and nutrition policymakers and managers from different disciplines for simplifying and aligning implementation materials avoids duplication of actions, optimizes resources, and motivates healthcare workers to comply with recommended priority health actions.
- Harmonized communication among health and nutrition approaches stimulates learning and supports healthy feeding and care behaviours, boosting the confidence of care providers and users.

Insights from the health system:

- Involving the MOH from the start through collaborative partnerships enhances strong leadership for context-adapted and strategic implementation.
- Local capacities are strengthened by building on the existing local health system, optimizing the use of limited resources and mitigating unintended negative consequences, such as competing or diluted efforts.
- Including a system for quality improvement and learning enhances implementation research to understand what works for whom under what circumstances and to improve practices.

Our collective active knowledge exchange unearthed insights and generated new knowledge, sparked by interactions within and across the teams. Co-learners valued understanding why and how intended effects happened or did not happen and applied it in “real time” to inform their own “what next?” Peer-to-peer learning facilitated by the international lead authors was non-judgmental and non-critical, which encouraged transparent open exchange.

Across all three settings, there was a longstanding felt need and demand for documenting learning. A committed “champion” was instrumental to initiate and navigate the approach. Our findings demonstrate how contextual realities and responses determine pathways to feasible care and how person-centred continuity of care delivered at integrated services are central to this.

To help bridge the know-do gap that impedes scalable care across nutrition and health, case studies are needed that not only describe what happened but also how. Implementation research requires pragmatic academic partners to maximize potential to evidence policy and make the most of practice innovations.

Implementing and support partners, such as WHO, UNICEF and NGOs, need to align their work with national health and nutrition policies, service planners, and implementation realities to improve readiness to embed and scale up a sustainable approach successfully within existing health services. Our case studies spotlight shortfalls and common constraints faced in routine service provision. To realize a pathway of care, there needs to be investment in routine service capabilities. The case studies also demonstrate that it is possible to be system supportive and scale sensitive in emergency responses. Given the protracted and repeated crises around the world, this is imperative.

## CONCLUSION

The MAMI Care Pathway approach was needed and feasible in the three diverse contexts. Health system strengthening and healthcare provider engagement are fundamental for impactful scale-up. Despite marked differences, the three settings shared commonalities that we can learn from. We hope that our “learning by doing, together” series contribute to global efforts to develop implementation guidance for managing at-risk infants u6m and their mothers (in all their shapes and forms) and national efforts to contextualize this guidance. We also hope that our series catalyze more in-depth case studies centered on local experience to significantly inform policy development and practice.

## Supporting information

Supplemental Material 2 Data Tools Sustainable Scale-Up

Supplemental Material 3 MAMI Care Pathway Unpacked

Supplemental Material 4 MAMI Care Pathway Criteria

Supplemental Material 2 Data Tools Planning for Implementation

Supplemental Material 2 Data Tools Planning for Scale-Up

Supplemental Material 2 Data Tools Normalization

## Data Availability

All data produced in the present study are available upon reasonable request to the authors.

## Notes

### Competing Interest Statement

The authors have declared no competing interest.

### Funding Statement

This research was funded by the Bill & Melinda Gates Foundation, and the Department of Foreign Affairs, Ireland.

### Summary of Updates

Author email and affiliation updated

## REFERENCES

1. Global Nutrition Report Stakeholder Group. 2021 Global Nutrition Report: The state of global nutrition. Bristol, UK; 2022.

2. Lawn JE, Ohuma EO, Bradley E, Idueta LS, Hazel E, Okwaraji YB, et al. Small babies, big risks: global estimates of prevalence and mortality for vulnerable newborns to accelerate change and improve counting. Lancet. 2023;401(10389):1707–19.

3. Marko Kerac PTJ, Marie McGrath, Eilise Brennan, Tim J Cole, Charles Opondo, and Severine Frison. Malnutrition in infants aged under 6 months: Prevalence and anthropometric assessment – secondary analysis of 56 low- and middle-income country DHS data. Under peer review. 2024.

4. Mertens A, Benjamin-Chung J, Colford JM, Coyle J, van der Laan MJ, Hubbard AE, et al. Causes and consequences of child growth failure in low- and middle-income countries. medRxiv. 2020:2020.06.09.20127100.

5. Victora CG, Christian P, Vidaletti LP, Gatica-Domínguez G, Menon P, Black RE. Revisiting maternal and child undernutrition in low-income and middle-income countries: Variable progress towards an unfinished agenda. The Lancet. 2021;397(10282):1388–99.

6. McGrath M, Wrottesley SV, Brennan E, Samnani A, Deconinck H. Invisible pursuit: global policy guidance on care of vulnerable infants under 6 months and their mothers, a scoping review. Emergency Nutrition Network (ENN), Kidlington, Oxford, UK. 2024.

7. Greenhalgh T, Papoutsi C. Studying complexity in health services research: desperately seeking an overdue paradigm shift. BMC Medicine. 2018;16(1):95.

8. Paparini S, Papoutsi C, Murdoch J, Green J, Petticrew M, Greenhalgh T, et al. Evaluating complex interventions in context: systematic, meta-narrative review of case study approaches. BMC Medical Research Methodology. 2021;21(1):225.

9. MAMI Global Network, ENN, LSHTM. MAMI Care Pathway Package. v3. Oxford, UK: Emergency Nutrition Network. https://www.ennonline.net/mamicarepathway; 2021.

10. Grey K, Brennan E, Kerac M, McGrath M. The MAMI Care Pathway Package: A resource to support the management of small and nutritionally at-risk infants under six months of age and their mothers (MAMI). South Sudan Medical Journal. 2021(14(3)):94–9.

11. World Health Organization. Guideline: updates on the management of severe acute malnutrition in infants and children. Geneva: World Health Organization; 2013.

12. World Health Organisation. WHO guideline on the prevention and management of wasting and nutritional oedema (acute malnutrition) in infants and children under 5 years. World Health Organisation. 2023.

13. World Health Organization. Integrated Management of Childhood Illness: Chart Booklet. Geneva, Switzerland: World Health Organization; 2014.

14. Devon Indig KL, Anne Grunseit, Andrew Milat, Adrian Bauman. Pathways for scaling up public health interventions. BMC Public Health. 2018.

15. Greenhalgh T, Papoutsi C. Studying complexity in health services research: desperately seeking an overdue paradigm shift. 2018;16:95.

16. Skivington K, Matthews L, Simpson SA, Craig P, Baird J, Blazeby J. A new framework for developing and evaluating complex interventions: update of Medical Research Council guidance. 2021;374:n2061.

17. World Health Organization, ExpandNet. Beginning with the end in mind: planning pilot projects and other programmatic research for successful scaling up. 2011.

18. MAMI Global Network [Internet]. Available from: https://www.ennonline.net/network/mami-global-network.

19. Pawson R. Evidence-based Policy: a Realist Perspective. London: Sage; 2006.

20. Project TRI. Retroduction in realist evaluation. The RAMESES II Project.

21. May CR, Cummings A, Girling M, Bracher M, Mair FS, May CM, et al. Using Normalization Process Theory in feasibility studies and process evaluations of complex healthcare interventions: a systematic review. Implement Sci. 2018;13(1):80.

22. Murray E, Treweek S, Pope C, MacFarlane A, Ballini L, Dowrick C. Normalisation process theory: a framework for developing, evaluating and implementing complex interventions. 2010;8:63.

23. Greenhalgh T, Wherton J, Papoutsi C, Lynch J, Hughes G, A’Court C, et al. Beyond Adoption: A New Framework for Theorizing and Evaluating Nonadoption, Abandonment, and Challenges to the Scale-Up, Spread, and Sustainability of Health and Care Technologies. J Med Internet Res. 2017;19(11):e367.

24. World Health Organization. Nine steps for developing a scaling up strategy. Geneva, Switzerland: World Health Organization; 2010.

25. World Population Review. Total population by country 2023: Pakistan: World Population Review; 2023 [Available from: https://worldpopulationreview.com/countries.

26. South Sudan National Bureau of Statistics. Projected Population 2020-2040 2023 [Available from: https://nbs.gov.ss/publications/south-sudan-population-projections-2020-2040/.

27. UNICEF. UNICEF Data: Monitoring the situation of children and women. 2023 [Available from: https://data.unicef.org/.

28. UNICEF. Policy brief - Improving quality of care around the time of birth in Pakistan. 2018.

29. National Institute of Population Studies (Pakistan) I. Pakistan Demographic and Health Survey 2017-18. 2019.

30. Organization WH. Trends in maternal mortality 2000 to 2017: estimates by WHO, UNICEF, UNFPA, World Bank Group and the United Nations Population Division. Geneva, Switzerland; 2019.

31. Group WB. 2023 [Available from: https://data.worldbank.org/.

32. Exclusive breastfeeding rates up in South Sudan as country marks, World Breastfeeding Week [press release]. 2019.

33. UNICEF. Scaling up care for children with severe acute malnutrition in South Sudan: lessons learned from expanding quality services in a complex emergency context. New York: UNICEF; 2020.

