## Supplemental Material 1 Methods for "Reflective Learning from Implementing Care Pathways for Vulnerable Infants and Their Mothers: Case studies from Pakistan, South Sudan and Yemen"

The case studies used mixed methods in which different theories applied different lenses to examine the introduction and implementation and the adoption of the MAMI Care Pathway in each case context and generate learning and ideas on improving implementation and scalability.

#### Inquiry Tools

**First**, a *Planning and Implementation Process Framework for the MAMI Care Pathway approach* was developed, inspired by the 2010 WHO ExpandNet 'Nine steps for developing a scaling-up strategy,' the 2011 WHO ExpandNet 'Beginning with the end in mind' (1) and tacit knowledge of co-researchers (Box A2.1). It was used to gain a detailed description of the planning and implementation process within the defined context of each country case.

**Box A2.1: Planning and Implementation Process Framework**

***Context***
- Country context
- Organisational context

***Situation analysis prior to starting***
- Burden and perceived health priority
- Policy context
- Local health system capacities
- Stakeholders

***Planning for implementation***
- Initiating discussions – Agency's preparedness
- Engaging key stakeholders
- Defining the target population
- Selecting sites for implementation
- Designing the implementation modus - tailoring the innovation to the local context and capacities
- Using, adapting, aligning, simplifying, testing materials
- Training for implementation

***Service delivery – implementation***
- Access: availability, geographic accessibility/delivery points, affordability, acceptability
- Organisation of care in the community, in the health facility
- Organisation of staff
- Participation
- Partnerships

***Monitoring, improving and collaborative learning***
- Monitoring and reporting
- Improving quality
- Disseminating information and learning
- Maintaining and sustaining quality services
- Ensuring accountability to users, managers and funders of the services
- Advocating for implementation and scale-up

***Suggestions for improving implementation***

**Second**, the *Normalization Process Theory* (NPT) provided a conceptual framework that helped to understand and evaluate the processes by which the MAMI Care Pathway approach was routinely operationalized in everyday work (2-4). The NPT used a participatory method to explore the four components of the adoption process to uncover what individuals and groups either do or do not do to enable normalisation of the intervention:

1) Coherence–meaning and sense-making–defines and organises the components of a practice;
2) Cognitive participation–commitment and engagement–defines and organises the people implicated in a complex intervention;
3) Collective action–work done to enable the intervention to happen–defines and organises the enacting of a practice; and
4) Reflective monitoring–reflect on or appraise the benefits–defines and organizes the assessment of the outcome of a practice.

The success of implementing the MAMI Care Pathway approach by health workers adopting the practice was scored by the case study team on a 5-point Likert sliding scale from 'not at all' (grade 1) to 'completely' (grade 5).

**Third**, the *Non-adoption, Abandonment, Scale-up, Spread and Sustainability* (NASSS) *Framework* was adapted and used in a participatory process to synthesise insights on evaluating adoption challenges that impact on scaling up and sustainability (5) (Figure A2.1). It was used as a reflexive guide to generate ideas on challenges related to: (1) the condition, (2) the technology, (3) the value proposition, (4) the adopters, (5) organisation, (6) the wider system, and (7) embedding and adapting over time. A grading system was used to express whether the challenges identified were simple, complicated, or complex: (1) simple meaning understandable or predictable, relatively straightforward to address; (2) complicated meaning less understandable, controllable, thus less straightforward to address; and (3) complex meaning not understandable or predictable, a dynamic or emergent behaviour.

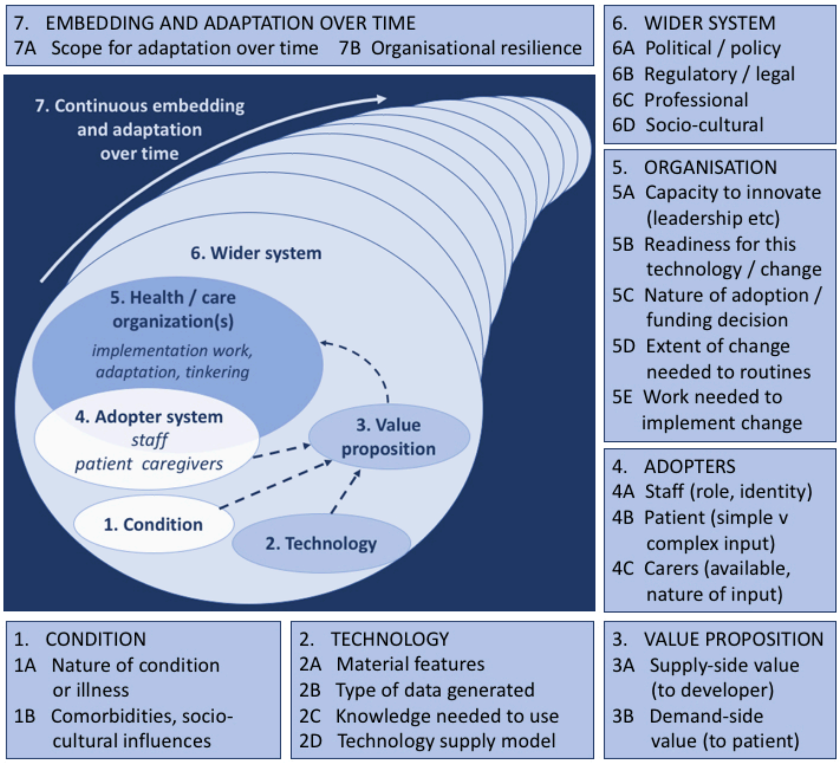


**Figure A2.1. The Non-adoption, Abandonment, Scale-up, Spread and Sustainability (NASSS) Framework for considering influences on the adoption, nonadoption, abandonment, spread, scale-up, and sustainability of a health intervention.**

**Fourth**, the *Checklist for Assessing the Potential Scalability* of pilot projects or research (1, 6) was used to explore how easy or difficult it would be to scale up each case and to provide insights into what steps to take to facilitate sustainable scale-up. The checklist provides recommendations in twelve steps on how to design pilot projects considering scale-up that lead to lasting and larger-scale impact (Box A2.2).

**Box A2.2. Twelve recommendations on how to design pilot projects with scaling up in mind**

| | |
|---|---|
| Step 1 | Engage in a participatory process involving key stakeholders |
| Step 2 | Ensure the relevance of the proposed innovation |
| Step 3 | Reach consensus on expectations for scale-up |
| Step 4 | Tailor the innovation to the sociocultural and institutional settings |
| Step 5 | Keep the innovation as simple as possible |
| Step 6 | Test the innovation in the variety of sociocultural and institutional settings where it will be scaled-up |
| Step 7 | Test the innovation under the routine operating conditions and existing resource constraints of the health system |
| Step 9 | Advocate with donors and other sources of funding for financial support beyond the pilot stage |
| Step 8 | Develop plans to assess and document the process of implementation |
| Step 10 | Prepare to advocate for necessary changes in policies, regulations and other health-systems components |
| Step 11 | Develop plans for how to promote learning and disseminate information |
| Step 12 | Plan on being cautious about initiating scale-up before the required evidence is available |

### Case Study Selection

Case selection sought a variety of implementation modalities or characteristics, e.g.:

- Implementing a care pathway addressing at-risk infants and their mothers, as a pilot, research or programme;
- Differences in terms of context, implementers, geography;
- Either government-led or partner-led;
- In a development, emergency or fragile setting;
- In a low- or middle-income country (LMIC) setting, either urban, rural or mixed;
- With availability of data on process and outcomes;
- With expressed interest and availability to participate in the case study;
- Either in an English or French speaking environment.

A primary selection criterion was that participating in this process would add value and contribute to local learning and progress on implementing the MAMI Care Pathway approach.

The country cases selected encompassed a variety of settings where the MAMI Care Pathway approach was applied:

- **Pakistan**: Paediatrician-led services in a private charity hospital in Karachi.
- **South Sudan**: Implementation study where the MAMI Care Pathway approach is integrated into maternal and child health services in urban and rural sites by the MOMENTUM Integrated Health Resilience project.
- **Yemen**: Pilot implementation integrated into a health and nutrition emergency programme by ADRA.

### Data Collection

An iterative and participatory process of reflective learning took place across four phases that built on each other. Data tools consisted of generic questionnaires that served as interview guides specifically developed for the MAMI Care Pathway approach and adapted to each country case (Box A2.3) (see Annex 3).

The first phase of investigation was largely descriptive through written feedback and clarification. Next, the shared information was built upon, through interviews, to further explore 'how' things happened or not, paying particular attention to social dimensions.

The second phase consisted of participatory discussions with clinical service providers which explored adoption of the MAMI Care Pathway approach as part of their routine work.

The third phase brought together senior managers and clinical health workers to discuss challenges in adopting the MAMI Care Pathway approach.

The fourth phase synthesised the discussion in the third phase across the country cases, allowing for reflection on potential scalability based on triangulating information collected across the three.

**Box: Data tools**

Phase 1: Questionnaire (written and oral investigation) using the Planning and Implementation Process Framework; respondents were (sub)national health, nutrition, MAMI managers or advisors.

Phase 2: Interview guide using the *Normalization Process Theory;* respondents were clinical healthcare workers implementing the care pathway approach.

Phase 3 and Phase 4a: Checklist for participatory group discussions using the *NASSS Framework;* respondents were the participating national and (sub)national health, nutrition, MAMI managers or advisors who discussed their country context in phase 3, and then came together to discuss across countries in phase 4a.

Phase 4b: *Checklist for Assessing the Potential Scalability* using the information generated across phases.

Respondents were asked to provide their informed consent prior to their participation and withdrawal from the inquiry was possible at any time.

Data was collected through written feedback and during interviews, which were digitally recorded following consent from all interviewees. Respondents could skip questions for any reason. Where possible, the reason for not answering was recorded but was not mandatory. Audio recordings were transcribed verbatim within 48 hours of collection using Otter.ai software. All digital data were stored in a password-protected digital space accessible only to investigators. All country-specific data was shared with the country teams.

During data collection and analysis, notes on possible biases, interferences or limitations were recorded and reported on.

### Analysis

*The stepwise and iterative inquiry* appraised the case study experiences by applying different lenses to generalise learning through repeated cycles of testing and building ideas (theories) of why things have worked or not and how (mechanisms of action). This 'theory-driven' iterative analysis involved the following steps:

> Descriptive data analysis: Data on introducing and implementing MAMI were summarised by topic to understand processes of planning, introducing, adapting, implementing, monitoring and improving the MAMI Care Pathway approach, to uncover what was done how, to appraise readiness for scale-up.
> Explorative data analysis: Data on perceptions of clinical healthcare workers on implementing and adopting the MAMI Care Pathway approach were analysed for emerging themes to explore perceptions on what worked for whom under what circumstances and appraised adoption.
> Explanatory data analysis: Data on descriptions and perceptions were triangulated and synthesised to inform updates and evolution of our theories/ideas on the MAMI Care Pathway approach and identify practical, pragmatic ways to help progress towards scalable, sustainable care.

Data was analysed both deductively (test our ideas/theories) and inductively (find new ideas/theories), involving the respondents and requesting their opinion, as well as confirming the generated ideas/theories. . Data was synthesised in each step by intuitive-reflective appraisal **–** which involved perceptions on what immediately felt right or made sense, and then questioning these by considering other possibilities.

*Participatory and adaptive, reflexive learning:* Interviewers and interviewees were involved in reflective learning building upon each step, thereby 'learning together by doing.' This collaborative 'learning together' deepened the understanding of embedding and adapting the MAMI Care Pathway approach in diverse local systems of health. Besides the strengthening of own capacities and understanding by tapping into implicit and often invisible and under-appreciated tacit knowledge, this approach was useful for contributing to overall collective learning on the 'how' of the MAMI Care Pathway approach.

### Limitations

Each country case covered introduction and implementation of the MAMI Care Pathway approach on a small scale in a specific context which limited the generalisability of learnings across broader systems and services within and across countries. Each case study also engaged a limited number of respondents (between two and four, depending on the case) which restricted the breadth of perceptions. However, the different lenses applied through the case study phases did generate an in-depth understanding for each case context, while identifying common theories/ideas which influence implementation, adoption, scale-up and sustainability, even across the diverse case contexts, thereby contributing to collective learning.

The qualitative approach involved online interviews which lack the human presence needed to build trust and to convey the subtleties of eye contact or body language which contribute to multidimensional and nuanced understanding of the ideas/perspectives shared (7).

Specifically, during Phase 2 (*interview guided by the Normalization Process Theory*), only one or two clinical health workers responsible for implementing the MAMI Care Pathway approach (assessment, support and progress monitoring of the mother-infant pair), were interviewed. The low numbers of people involved likely limited the perceptions on the normalization process. The clinical health worker responding was also either an existing, or a newly recruited, staff member accompanied by a trained supervisor or assistant which may have influenced their answers. Responses often fell into discussions on 'perceived benefits' of the MAMI Care Pathway approach, rather than building on perceptions of the adoption process. Finally, discussions went in various directions, and sometimes the same elements were repeated, or questions were not answered well, or the answer fitted a question that would come later. This resulted in some reorganisation of responses to fit the flow of the interview guide after the discussion.

### REFERENCES


1. World Health Organization, ExpandNet. Beginning with the end in mind: planning pilot projects and other programmatic research for successful scaling up. 2011.
2. May CR, Finch T, Ballini L, MacFarlane A, Mair F, Murray E, et al. Evaluating complex interventions and health technologies using normalization process theory: development of a simplified approach and web-enabled toolkit. BMC Health Services Research. 2011;11(1):245.
3. Murray E, Treweek S, Pope C, MacFarlane A, Ballini L, Dowrick C, et al. Normalisation process theory: a framework for developing, evaluating and implementing complex interventions. BMC Medicine. 2010;8(1):63.
4. May C, Rapley T, Mair FS, Treweek S, Murray E, Ballini L, et al. Normalization Process Theory on-line users' manual, toolkit and NoMAD instrument 2015.
5. Greenhalgh T, Wherton J, Papoutsi C, Lynch J, Hughes G, A'Court C, et al. Beyond adoption: A new framework for theorizing and evaluating nonadoption, abandonment, and challenges to the scale-Up, spread, and sustainability of health and care technologies. J Med Internet Res. 2017;19(11):e367.
6. World Health Organization, ExpandNet. Nine steps for developing a scaling-up strategy. 2010.
7. You S. Feminist Perspectives [Internet]: King's College London. 2021. Available from: https://www.kcl.ac.uk/challenges-and-gains-in-conducting-online-interviews-during-the-pandemic.
