## Supplemental Material 2 Data Tools Sustainable Scale-Up for "Reflective Learning from Implementing Care Pathways for Vulnerable Infants and Their Mothers: Case studies from Pakistan, South Sudan and Yemen"

### Data Tool – Scale-up, Spread and Sustainability of the MAMI Care Pathway Approach

Applying the (non)adoption, abandonment, scale-up, spread, and sustainability (NASSS) framework in real time (Greenhalgh et al., 2017).

| | | | |
|---|---|---|---|
| **Respondents** | | | |
| **Date of interview** | | | |
| **Context (**where, since how long, whom, purpose/design) | | | |
| **ORIGINAL NASSS QUESTIONS** | **ADAPTED NASSS QUESTIONS** | **GRADING CONSIDERATIONS**<br>1= understandable or predictable aspects are relatively straightforward to address (simple).<br>2= less understandable or predictable aspects or many factors are involved (complicated).<br>3= inherently not understandable or predictable, but dynamic or emergent aspects are involved(complex). | **RESPONSE** |
| **Domain 1: The condition or illness (risk factors)**<br>Addresses how well the **condition** 'small and nutritionally at-risk infants and their mothers' is a) characterized, understood, predictable, and b) how care is being affected by sociocultural factors and comorbidities. | | | |
| 1a. What is the nature of the condition or illness? | 1a. Is the condition 'small and nutritionally at-risk infants and their mothers' well-characterised, well-understood, predictable? | 1–Is the condition well characterized, well-understood, predictable? OR 2–Not fully characterized, understood or predictable? OR 3–Poorly characterized understood, unpredictable? | |
| 1b. What are the relevant sociocultural factors and comorbidities? | 1b. Are sociocultural factors and comorbidities relevant for the condition 'small and nutritionally at-risk infants and their mothers'? | 1–Are sociocultural factors and comorbidities unlikely to affect care significantly? OR 2–Affect care and must be factored in? OR 3–Pose significant challenges to care planning and service provision? | |
| **Domain 2: The technology**<br>Addresses whether **the methods (technologies)** of the MAMI Care Pathway used for detecting, classifying, and supporting 'small and nutritionally at-risk infants and their mothers' are a) newly introduced, b) need new knowledge, c) need continued support, and d) need specific adaptations. | | | |
| 2a. What are the key features of the technology? | 2a. What are key features of the methods (technologies) to assess, classify, support 'small and nutritionally at-risk infants and their mothers'? Are methods known, do they exist? | 1–Are methods (technologies) to assess, classify, support 'small and nutritionally at-risk infants and their mothers' already installed or existing, dependable? OR 2–Are they new to develop? OR 3–Do they need to be embedded in an existing (complex) system? | |
| 2b. What kind of knowledge does the technology bring into play? | 2b. Is new knowledge generated or made visible when applying the methods to assess, classify, support 'small and nutritionally at-risk infants and their mothers'? Has it the potential to detect changes in health and nutrition status? | 1–Do the methods to detect, classify, support 'small and nutritionally at-risk infants and their mothers' make risks or changes in risks visible or measurable? OR 2–partially or indirectly visible/measurable? OR 3–Changes are unpredictable or can be contested? | |
| 2c. What knowledge and/or support is required to use the technology? | 2c. What knowledge and/or technical support is required to assess, classify, support 'small and nutritionally at-risk infants and their mothers'? | 1–No new knowledge is required to assess, classify, support 'small and nutritionally at-risk infants and their mothers'? OR 2–Detailed instructions and training are needed? OR 3– Advanced training and support are necessary? | |
| 2d. What is the technology supply model? | 2d. Are the methods used in the MAMI Care Pathway generic, standardised? | 1–Are the 'small and nutritionally at-risk infants and their mothers' methods used in the approach generic, standardised, straightforward to implement? OR 2–Are significant organizational changes in the management of health services needed? OR 3–Is it highly vulnerable to support withdrawal? | |
| **Domain 3: The value proposition**<br>Explores whether the MAMI Care Pathway is considered a **valuable intervention and for who** it has value, a) the care provider and b) the user. | | | |
| 3a. What is the developer's business case for the | 3a. How do health workers (HWs) perceive the value of the MAMI Care Pathway? Do they understand | 1–Is the perceived benefit of the MAMI Care Pathway approach well understood, on the short/mid/long term? OR 2–Is it undervalued (at risk?) OR 3–Is it | |

| | | |
|---|---|---|
| technology (supply-side value)? | the value of the short/mid/long term benefits? | unlikely that it will be maintained (after the pilot period), at risk? |
| 3b. What is its desirability, efficacy, safety, and cost effectiveness (demand-side value)? | 3b. How do the mothers (caregivers) perceive the value of the MAMI Care Pathway? Do they understand the need, do they appreciate the care, is the opportunity cost a barrier? | 1–Is the MAMI Care Pathway approach considered needed, desirable, safe, cost-effective by the user? OR 2–Is it unknown, contested? OR 3–Is it considered not needed, undesirable, unsafe, ineffective or unaffordable by the user? |
| **Domain 4: The adopter system**<br>Explores whether the MAMI intervention has been **adopted (accepted) and by who:** a) health staff, b) mothers, c) lay support system of the mother. | | |
| 4a. What changes in staff roles, practices, and identities are implied? | 4a. Did important changes have to be made for health workers (staff in the health facility) to take on their role in the MAMI Care Pathway? Did new skills have to be learned, new staff be appointed, new tasks to be taken on? | 1–When adopting the care pathway, were there no changes in staff roles and practices? OR 2–Did existing staff have to learn new skills and/or was new staff appointed? OR 3–Posed it a threat to current professional identities, values and scope of practices (risk of job loss)? |
| 4b. What is expected of the patient (and/or immediate caregiver)—and is this achievable by, and acceptable to, them? | 4b. Were specific or new actions expected of the mother? | 1–Nothing is expected of the mother (principal caregiver). OR 2–Routine tasks and changes in behaviour are expected. OR 3–Complex tasks are expected, and are these achievable, acceptable? |
| 4c. What is assumed about the extended network of lay caregivers? | 4c. By offering MAMI, are other lay care givers in the mother's network affected (e.g., family members, volunteers, community members), and are there new requirements or expectations for them? Is the wider network requested to involve? | 1–Nothing is required from the extended network of lay caregivers. OR 2–Caregivers are assumed to be available. OR 3–A network of caregivers is needed/expected to coordinate their inputs. |
| **Domain 5: The organization**<br>Addresses whether **the organisation** of the MAMI intervention required important changes and inputs in the given organizational context: a) capacity, b) readiness to adopt, c) easiness of adoption and funding decision, d) changes in teamwork, and e) tasks to be undertaken (the work). | | |
| 5a. What is the organization's capacity to innovate? | 5a. Did the organizational set-up had the capacity to innovate, change, adapt ways of working, and have the resources for doing so? | 1–Local health system is well organized (good managerial capacity, well supported), flexible available resources, good management, risk taking is encouraged. OR 2–Resources are inflexible, local leadership is suboptimal and risk taking is not encouraged. OR 3–Severe resource pressure, weak leadership, weak resilience. |
| 5b. How ready is the organization for this technology-supported change? | 5b. Was the organizational set-up ready / open to innovate, change, adapt ways of working, and have the resources for doing so? | 1–High tension for change, openness for innovation, widespread support. OR 2–Little tension for change, moderate innovation. OR 3–No tension for change, poor innovation, opponents to change. |
| 5c. How easy will the adoption and funding decision be? | 5c. How easy will the adoption and funding decision for the MAMI Care Pathway be (resources, cost savings, new infrastructure to manage by the MOH, NGO or donor lead)? | 1–Single organization with sufficient resources; anticipated cost savings; no new infrastructure or recurrent costs required. OR 2–Multiple organizations with partnership relationship; cost-benefit balance favourable or neutral; new infrastructure found (e.g., repurposing staff roles, training). OR 3–Multiple organizations with no formal links and/or conflicting agendas; funding depends on cost savings across system; costs and benefits unclear; new infrastructure conflicts with existing; significant budget implications. |
| 5d. What changes will be needed in team interactions and routines? | 5d. What changes were needed in MOH, NGO, health worker team organization to adopt MAMI? Did team interactions and team | 1–No new team routines or care pathways needed. OR 2–New team routines or care pathways that align readily with existing ones. OR 3–New team routines or care pathways that conflict with existing ones. |

| | | |
|---|---|---|
| | routines change (new), align or conflict? | |
| 5E. What work is involved in implementation and who will do it? | 5e. What work is involved in implementing and improving the quality, and who will do it? | 1–Established shared vision, few simple tasks, uncontested and easily monitored. OR 2–Some work needed to build shared vision, engage staff, enact new practices, monitor impact. OR 3–Significant work needed to build shared vision, engage staff, enact new practices, monitor impact. |
| **Domain 6: The wider context**<br>Explores whether **financial and policy requirements** are in place nationally for roll out. | | |
| 6a. What is the political, economic, regulatory, professional (e.g., medicolegal), and sociocultural context for programme rollout? | 6a. Are financial and policy requirements for MAMI in place for programme roll out? a/ the past context, b/ the future context for expansion? | 1–Financial and regulatory requirements are in place nationally; professional bodies and civil society are supportive. OR 2–Are being negotiated nationally; professional bodies and lay stakeholders not yet committed. OR 3–Raise tricky or legal or other challenges, professional bodies and lay stakeholders are opposed. |
| **Domain 7: Embedding and adaptation over time**<br>Explores the feasibility of **embedding and adapting** the MAMI approach over time: feasibility of a) continuing to adapt and evolve on the medium and long-term, and b) building organisational resilience. | | |
| 7a. How much scope is there for adapting and coevolving the technology and the service over time? | 7a. What is the feasibility of continuing embedding and adapting the MAMI approach (intervention modalities) over time (medium-long term)? Are you expecting certain barriers? | 1–Strong scope for adapting and embedding the MAMI approach. OR 2–Potential for adapting and coevolving the MAMI services are limited and uncertain. OR 3–Significant barriers to the further adaptation or co-evolution of the MAMI approach. |
| 7b. How resilient is the organization to handling critical events and adapting to unforeseen eventualities? | 7b. What is the organisational resilience to detect and overcome critical issues or barriers (barriers related to embedding, handling critical events, adapting to unforeseen eventualities?) | 1–Sense-making, collective reflection and adaptive action are ongoing and encouraged. OR 2–Are difficult and viewed as low priority. OR 3–Are discouraged in a rigid, inflexible implementation model. |
