## Supplemental Material 3 MAMI Care Pathway Unpacked for "Reflective Learning from Implementing Care Pathways for Vulnerable Infants and Their Mothers: Case studies from Pakistan, South Sudan and Yemen"

**Table 1: MAMI Care Pathway activities unpacked, Pakistan, South Sudan and Yemen cases, 2023**

| | | | Pakistan case | South Sudan case | Yemen case |
|---|---|---|---|---|---|
| **Activity** | **Detailed activity** | **What** | | **Where and by who** | |
| **Community:** | | | | | |
| **Sensitization** | **Sensitization creating awareness of risks and participation in care** | Sensitize and discuss risks of poor growth and development of infant, risks related to the mother, and the MAMI Care Pathway with community members through, for example, community activities, mother groups, household visits. | NA | Household visits, social gatherings<br>By CHW | Household visits, social gatherings<br>By CHW, community midwives |
| **Screening** | **Community screening** | Discuss with mothers to verify key indicators, and referral to health centres in case of identified risk. | | | |
| **Follow-up** | **Community monitoring and counselling of at-risk pairs** | Follow up and counsel at-risk pairs through household visits; find pairs that default or miss follow-up. | | | |
| **Health facility:** | | | | | |
| **Sensitization** | **Sensitization creating awareness of risks and participation in care** | Sensitize and discuss risks of poor growth and development of infant, risks related to the mother, and the MAMI Care Pathway with health facility attendants | Hospital (MAMI clinic, inpatient department, waiting areas)<br>By doctors and nurses | PHC registration and waiting areas<br>By HCW | PHC registration and waiting areas<br>By trained HCW |
| **Health promotion** | **Health and nutrition education** | Provide general health and nutrition education on essential family practices and prepare for quality complementary feeding. | Hospital MAMI clinic, pediatric OPD<br>By doctors and nurses | PHC registration and waiting areas<br>By trained HCW | PHC registration and waiting areas<br>By trained HCW |

| | | | | | |
|---|---|---|---|---|---|
| **Screening** | **Mother and child health services screening of all mother-infant pairs** | Ask mothers questions to verify key indicators and measure MUAC of infants and mothers. | Hospital MAMI clinic<br>By doctors and nurses | Relevant PHC units (immunization, maternity, antenatal/postnatal care, nutrition, family planning, under-five clinic)<br>By trained HCW | Relevant PHC units<br>By trained HCW |
| **Assessment** | **Anthropometric assessment** | Measure anthropometry of screened infant–mother pairs identified at-risk: weight, length, MUAC, WAZ, oedema check. | | PHC registration, waiting area or nutrition unit<br>By trained HCW | PHC OPD nutrition unit<br>By trained HCW |
| | **IMNCI risk assessment**<br>**Feeding assessment** | Conduct a comprehensive IMNCI and MAMI risk assessment. | | PHC OPD or under-five clinic<br>By clinical HCW | |
| | **Mental health assessment** | | Hospital Psychosocial department<br>By psychologist | | |
| | **Classification of risk** | Classify risk and decide on enrolment or referral. | Hospital MAMI clinic<br>By doctors and nurses | | |
| **Outpatient support** | **Enrolment** | Register and copy information from the assessment as baseline. | | PHC OPD or under-five clinic<br>By clinical HCW | PHC OPD nutrition unit<br>By trained HCW |
| | **Initial targeted counselling** | Counsel on identified concern related to ill health, feeding difficulty of infant | | | |
| | | Counsel on identified problems related to mental health concern of mother. | Psycho-social department<br>By psychologist | | |
| | **Follow-up monitoring** | Repeat assessment, monitor progress and refer if needed. | Hospital MAMI clinic | | |

| | | |
|---|---|---|
| | | By doctors and nurses |
| **Follow-up targeted counselling** | Counsel on previous or newly identified ill health, feeding difficulty, ECD concern of infant. | |
| | Counsel on previous or newly identified mental health concern of mother | Psycho-social department<br>By psychologist |
| **End of care when infant reaches six months of age** | Review outcome and decide on referral to follow-on services. | |

CHW= community health worker; ECD= early childhood development; HCW= healthcare worker; MAMI= Management of small and nutritionally at-risk infants under 6 months of age and their mothers; NA= not applicable; OPD= outpatient department; PHC=primary healthcare centre; WAZ= weight-for-age z-score; WLZ= weight-for-length z-score
