## Supplemental Material 4 MAMI Care Pathway Criteria for "Reflective Learning from Implementing Care Pathways for Vulnerable Infants and Their Mothers: Case studies from Pakistan, South Sudan and Yemen"

**Table 2: Criteria to identify infants u6m and their mothers in the MAMI care pathway, Pakistan, South Sudan and Yemen cases, 2023**

| Pakistan case | South Sudan case | Yemen case |
|---|---|---|
| **Criteria used to identify at-risk infants u6m and their mothers during rapid screening:** | | |
| ***Infant u6m:*** | | |
| • LBW<br>• Preterm birth (<37 weeks)<br>• Small for gestational age<br>• Mixed feeding or top-up feeding | • Small or LBW<br>• Clinically unwell, current illness<br>• Feeding difficulty<br>• Not breastfed<br>• Recent weight loss or failure to grow<br>• MUAC <125 mm | • LBW<br>• Preterm birth<br>• MUAC <110 mm (infant <6 weeks)<br>• MUAC <115 mm (infant ≥6 weeks)<br>• Excessive crying or sleep problems, colic and other concerns |
| ***Mother of infant u6m:*** | | |
| • Lactation difficulty<br>• MUAC <230 mm<br>• Mental health concern | • Absent or dead<br>• Ill health<br>• Multiple birth<br>• Adolescent mother <19 years<br>• MUAC <230 mm<br>• Mental health concern<br>• Other wellbeing or social concern | • Absent or dead<br>• Ill health<br>• Multiple birth<br>• Adolescent mother <18 years<br>• MUAC <230 mm<br>• Mental health concern |
| **Criteria used to identify moderate-risk infants u6m and their mothers during in-depth assessment:** | | |
| ***Infant u6m:*** | | |
| • LBW<br>• Preterm<br>• Small for gestational age<br>• Feeding difficulty<br>• Mixed feeding or top-up feeding<br>• MUAC ≥110 and <115 mm<br>• WAZ ≥–3 and <–2 or LAZ ≥–3 and <–2 | • (If known) LBW<br>• (If known) preterm<br>• Recent weight loss, no weight gain, poor growth<br>• Feeding difficulty<br>• Not breastfed<br>• MUAC <110 mm (infant <7 weeks)<br>• MUAC <115 mm (infant ≥7 weeks)<br>• WAZ <–2<br>• Excessive crying, sleep problems<br>• Other health concern or disability | • LBW<br>• Preterm<br>• Recent weight loss, no weight gain, poor growth<br>• Feeding difficulty<br>• MUAC <110 mm (infant <6 weeks)<br>• MUAC <115 mm (infant ≥6 weeks)<br>• WLZ <–2 and ≥–3<br>• WAZ <–2 and ≥–3<br>• Excessive crying, sleep problems<br>• Other health concern |
| ***Mother of infant u6m:*** | | |
| • Lactation difficulty<br>• MUAC <230 mm<br>• Mental health concern (PHQ-9) | • Absent or dead<br>• Multiple birth<br>• First child<br>• Adolescent mother <19 years<br>• Lactation difficulty | • Absent or dead<br>• Multiple birth<br>• Adolescent mother <18 years<br>• MUAC <230 mm<br>• Ill health |

| | | |
|---|---|---|
| | • MUAC <230 mm<br>• Ill health<br>• Moderate mental health concern (PHQ-9)<br>• Disability impairing feeding and/or care<br>• Other health or social concern | • Moderate mental health concern (PHQ-9) |
| **Criteria used to identify high-risk infants u6m and their mothers during in-depth assessment:** | | |
| ***Infant u6m:*** | | |
| • Infant with confirmed medical condition (e.g., congenital heart disease, metabolic disease, cerebral palsy)<br>• MUAC <110 mm<br>• WAZ <–3 or LAZ <–3<br>• Infant with weight loss at follow-up | • IMNCI general danger signs or signs and symptoms of severe disease, including nutritional oedema | • IMNCI general danger sign or sign and symptom of severe disease, including nutritional oedema<br>• WLZ <–3 or WAZ <–3 |
| ***Mother of infant u6m:*** | | |
| • Depression identified by a psychologist<br>• Persistent MUAC <230 mm after management<br>• Persistent anemia after management | • Severe physical or mental health concern (PHQ-9) | • Severe mental health concern (PHQ-9) |

<= below; ≥ = equal or above; IMNCI= Integrated management of neonatal and childhood illness: MUAC= mid-upper arm circumference; LAZ= length-for-age z-score; LBW= low birth weight <2500 g; PHQ-9= Patient Health Questionnaire-9 mental health score; WAZ= weight-for-age z-score; WLZ= weight-for-length z-score.
