## Supplemental Material 2 Data Tools Planning for Implementation for "Reflective Learning from Implementing Care Pathways for Vulnerable Infants and Their Mothers: Case studies from Pakistan, South Sudan and Yemen"

### Data Tool – Planning for Implementing the MAMI Care Pathway Approach

*[Note that the questions in green colour are discussed orally, all others are dealt with in writing.]*

**Responder(s)** *(name and function):* ________________

***Date of response:*** ________________

**Agency:** ________________

#### *1. Context*

##### 1.1 Country context relevant to MAMI

1. Describe the demographic and socio-economic context of your country, or the area where you are active.
   (E.g., development or emergency context, stable or fragile/fast changing/chronic, demographic pressure, climate change, political instability or insecurity, rural versus urban population, poverty, migration trends)

2. Describe key determinants that define vulnerability in infants under six months of age (u6m) and young children (data from the most recent survey/surveillance).
   (E.g., exclusive breastfeeding rate, inappropriate/harmful feeding and care practices, adolescent mothers, low birth weight)

##### 1.2 Organisational context for starting MAMI

3. Give name of agency or programme, and a brief description.
   (E.g., expertise/mandate, aim, activities, period of interventions, impact area, future plans, donor)

4. Give the justification for starting MAMI.
   (E.g., expected change, added value, opportunity, contribution, the MAMI Care Pathway could bring)

5. Explain who or what was the tipping point for deciding to start MAMI.
   (E.g., what or who was driving, motivating, enabling the decision; who or what enabled it just then and not earlier)

6. Give the aim or objective of the MAMI project that was defined at the start (and expected result if stated).

#### *2. Situation analysis prior to starting MAMI*

##### 2.1 Burden and perceived health priority

7. Give national key health and nutrition indicators (and trend) (with source and year, most recent survey, surveillance). Use the example table to answer.

Example table: Health and demographic information

| Information (Year, Source) | Data |
|---|---|
| Population (YYYY, Ref) | |
| Population at the MAMI sites YYYY, Ref) | |
| Fertility rate (YYYY, Ref) | |
| Live birth rate YYYY, Ref) | |
| Neonatal mortality YYYY, Ref) | |
| Infant mortality (YYYY, Ref) | |
| Low birth weight (YYYY, Ref) | |
| Assisted deliveries (YYYY, Ref) | |
| Exclusive breastfeeding YYYY, Ref) | |
| Global acute malnutrition 6-59m YYYY, Ref) | |
| Trend information (YYYY, Ref): | |

8. Prior to introducing MAMI, was the "vulnerability in infants u6m" recognised as a health or nutrition priority? Specify why or why not, by whom (in your opinion).
   (E.g., for the Ministry of Health (MOH) not a priority presuming that the needs are covered by the various policies and services; for [Agency] a priority because of deteriorating indicators in their impact area)

##### 2.2 Policy context

9. Did you do a policy analysis prior to starting MAMI?

10. If yes, describe what you did, scope, which tool you used. Use the example table to answer.
(E.g., national integrated management of acute malnutrition (IMAM) guideline covers inpatient treatment of wasting based on weight-for-height z-score (WHZ) <-3 z-score and presence of nutritional oedema in infants u6m; community infant and young child nutrition (IYCN) strategy advises to assess breastfeeding problems and counsel or refer during community growth monitoring sessions; guidelines on mental health cover post-partum depression; guidelines on small and sick newborns include targeted counselling)

Example table: Health and nutrition policy covering infants u6m and their mothers

| Policy, guideline (title, year) | Defined vulnerability in infants u6m and their mothers | Proposed interventions |
|---|---|---|
| xx | Xx | Xx |

- If no, why not?

##### 2.3 Local health system capacity

11. Did you do a capacity analysis/implementation readiness of the local health system or a feasibility study prior to starting MAMI (or any quick appraisal of readiness of the health facilities that involve in MAMI)?
    - If yes, describe what you did, which tool you used, when you did it in regard to starting MAMI, what are the headlines on what you found.
    - If no, why not?

12. List which MAMI activities were already covered at the community, primary care and tertiary care levels in the planned MAMI sites that were identified prior to starting MAMI?
(E.g., counselling on breastfeeding difficulties is done by nutrition assistants in the health centre and by community health workers and volunteers in the community as part of the national IYCN strategy)

13. List gaps in services, care, referral for infants u6m and their mothers that were identified prior to starting MAMI?

##### 2.4 Stakeholders

14. Did you do a stakeholder analysis prior to starting MAMI (quick appraisal of who is a MAMI stakeholder, and how to solicit their interest for involving early for what)?
    - If yes, describe what you did, which tool you used, when you did it in regard to starting MAMI, what are headlines on what you found. Please share any report on findings.
    - If no, why not?

15. Could you identify who is a relevant current or future stakeholder to involve in the design, planning, implementation; list who and specify why?

16. Did (could) you identify potential MAMI champions able to generate political will? If yes, who are they?
*(Note: a champion is an influential person who promotes 'a topic' and inspires others to take a more active role in that topic.)*

17. List key stakeholders you contacted and had preliminary discussions with on, e.g., introducing MAMI, sharing plans, probing their interest to be involved. Use the example table to answer.
(E.g., MOH Community Health Department – ways of strengthening active case finding of vulnerable infant-mother pairs, as part of existing community services)

Example table: Level of interest of key stakeholders to involve in MAMI

| Agency, department | Discussion topics on MAMI and level of interest | Name and email contact if appropriate |
|---|---|---|
| xx | xx | xx |

#### *3. Planning for MAMI implementation*

18. Give an indicative time line (# months) for inception discussions, designing and planning.

##### 3.1 Initiating discussions - Agency's preparedness

19. Describe key elements of the initial discussions and steps your agency undertook internally, prior to deciding and planning for MAMI implementation.
(E.g., internal discussion and decision, securing funds for which time span from which source–part of ongoing project, cost extension, additional budget–, hiring staff, securing equipment, planning)

20. Describe key elements of the initial discussions and steps your agency undertook externally, prior to deciding and planning for MAMI implementation.
(E.g., contacted MOH to discuss the relevance or perceived need, explore their interest in the innovation, feasibility, alignment or integration into the country's health system, roles and responsibilities, departments and technical partners to involve)

21. From whom did you seek approval for introducing MAMI, and how was this approval granted or formalised?

22. Was there a request for a formal description of the project prior to starting? If yes, describe the process, involvement of stakeholders and timeline.
(E.g., a project outline was shared and reviewed and approved by the MOH, taking two weeks; a study protocol was developed in participation with the MOH and approved (no IRB) taking two months)

23. Did you consult professional expertise within your agency; did you seek support externally? If yes, give profile of expertise and timeline.

- Did your agency conduct formative research prior to starting MAMI, or did you use in-house formative research? If yes, what? Share any reports.
(Note: formative research typically is done before starting a programme to understand practices and behaviours, needs for an intervention, e.g., a knowledge, attitudes, practices (KAP) survey for a reproductive health project)

**3.2 Engaging key stakeholders in the planning process**

24. Did you engage with the national and/or local MOH for planning the integration/implementation? Explain how and on what.

25. Who else you engaged with? Explain how and on what.
(E.g., UNICEF in face-to-face meeting and orientation workshop, for planning and review of materials, offering support for training as resources persons, offering scales and MUAC tapes)

26. In case you organised a meeting or workshop, describe who (and number) participated, how many days, what was the objective and outcome, what topics were covered, what documentation was shared.

27. Did key health and nutrition actors perceive MAMI a relevant innovation? Explain why or why not.

28. Are there lessons you want to share about the process?

**3.3 Defining the target population**

29. What criteria have been used to define vulnerability in infants u6m, and their mothers?

30. How were key health and nutrition actors involved in defining the target population for MAMI?

31. Are there lessons you want to share about the process?

**3.4 Selecting sites for implementation**

32. How did you define a MAMI implementation site in your project?
(E.g., specify the type of health facilities selected for implementing the outpatient Care Pathway, whether referral sites for inpatient care are involved, whether communities in the health catchment area covered, whether links between different sectors at different levels are established)

33. What criteria were used to select the sites?
(E.g., agency-supported health facilities; referral hospital with inpatient care for severe acute malnutrition)

34. Did key health and nutrition actors involve in selecting the sites? Explain.

35. Are there lessons you want to share about the process?

**3.5 Designing the implementation modus**

36. Did you tailor the implementation design for MAMI to the local context and capacities? If yes, explain how you did this, with whom and with what tools (if any)?

(E.g., participatory discussions with key stakeholders in a meeting using the 'who what where map'; informal discussion amongst agency staff)

37. Did you foresee ways of testing and/or adapting the implementation modus based on learning and feedback?

38. How did you appraise the capacity for absorbing MAMI by the local health system, at the selected health facility sites prior to implementing? What tools did you use, what difficulties did you anticipate, how did you plan to fill the gaps?
(E.g., consider gaps in knowledge, skilled health workers, equipment, space, referral services)

39. Are there lessons you want to share about the process?

##### 3.6 Using, adapting, aligning, simplifying, testing materials

40. Did you use and/or adapt the MAMI Care Pathway v3 materials? If yes, list which of the v3 materials were adapted and how this was done. Use the example table to answer.

Example table: Adaptation of MAMI Care Pathway v3 materials

| MAMI Care Pathway v3 material adapted | Description of adaptation(s) (what) | Method (how) |
|---|---|---|
| X | xx | Xx |
| X | xx | Xx |

41. Did you use existing materials for use in the MAMI Care Pathway? Use the example table to answer.

Example table: Existing materials used and/or adapted in MAMI

| Other materials used (adapted) | Description (what) | Method (how) |
|---|---|---|
| X | xx | Xx |
| X | xx | Xx |

42. Did you develop additional materials? Use the example table to answer.

Example table: Materials developed for use in MAMI

| Materials developed for use | Description (what) | Method (how) |
|---|---|---|
| X | xx | Xx |
| X | xx | Xx |

43. Who was involved in deciding the final version of materials to use?

44. Did you test the adapted materials prior to using them for implementation? If yes, describe how this was done.

45. Which (if any) materials were translated in a local language?

46. Describe how you overcame the local language barrier.
(E.g., developed a local language vocabulary as a cheat sheet and field tested it).

47. What were key challenges in the adaptation process?

48. Are there lessons you want to share about the process?

##### 3.7 Training for implementation

49. Did you train health workers ahead of implementing MAMI? If yes, explain who was trained (participants), on what (topics), by whom (trainers), how (method), with what materials, for how long (number of days), aiming to achieve what (learning objectives). Use the example table to answer.

Example table: Training for MAMI prior to starting

| Training (type and dates) | Participants targeted (profile and #) | Topics covered | Materials used | Learning objectives |
|---|---|---|---|---|
| Xx | xx | xx | xx | xx |

50. Were the national and/or local MOH involved in training? If yes, explain.

51. Were supervisors and managers involved in training? If yes, explain.

52. Were existing national or global training materials used? If yes, explain.

(E.g., on breastfeeding, IMNCI, counselling)

53. Did the training develop specific skills? If yes, explain.
(E.g., on using the IMNCI approach, measuring anthropometry, assessing breastfeeding, assessing mental health, targeted counselling)?

54. What skills were considered pre-requisite (skills training not covered)?

55. If you used the MAMI Care Pathway v3 materials, describe how you used these for training.

56. Are there lessons you want to share about the process?

#### 4. *Service delivery – implementation*

*[Notes:*
***Health services delivery** is about how services are organised and managed to ensure access, quality, safety, and continuity of care across health conditions across different locations and over time. Its core principles are:*

> *Comprehensive, equitable, sustainable, coordinated, continuous, holistic, preventive, empowering, goal oriented, respectful, collaborative, co-produced, endowed with rights and responsibilities, shared accountability, evidence-informed, led by whole-systems thinking, ethical.*

***People-centred care** is an approach to care that consciously adopts the perspectives of individuals, carers, families and communities as participants in, and beneficiaries of, trusted health systems that respond to their needs and preferences in humane and holistic ways. People-centred care also requires that people have the education and support they need to make decisions and participate in their own care.*

*https://apps.who.int/iris/bitstream/handle/10665/155002/WHO_HIS_SDS_2015.6_eng.pdf?sequence=1&isAllowed=y]*

57. Give an indicative time line for starting implementation support (enrolling first pair).

58. Give an indicative time line (# months) for ending implementation support (exiting of last pair, if relevant).

##### 4.1 Access: availability, geographic accessibility/delivery points, affordability, acceptability

59. Specify the geographical area and sites where MAMI is implemented. Use the example table to answer.
(E.g., region, districts, health facilities, start/end date)

Example table: MAMI sites

| Region | Health district | Primary care health centre | Referral hospital |
|---|---|---|---|
| Total | | | |

60. Did implementation start at all sites at the same time? If not, why not, how then?

61. Are services free of cost for small vulnerable infants and their mothers? Explain

62. If referral is needed, who organises, who pays for transport? Explain.

63. If referral for inpatient care is needed, who pays the admission fee, who pays for food for the caregiver? Explain.

64. Has your agency plans to expand or scale up MAMI in-country? In other countries? Specify what actions would facilitate this move?

##### 4.2 Organisation of care in the community (evidence-based, continuity (referral), coordinated, integrated, comprehensive, people-centred, equipped, equity)

65. What activities are provided at the community, how, where by whom? Use the example table to answer.

Example table: Who delivers where what services in the community

| Activities | How | Where | By whom |
|---|---|---|---|
| Sensitization | | | |
| Health and nutrition promotion | | | |
| Screening | | | |
| Referral | | | |
| Follow-up in the home during enrolment | | | |

66. Which MAMI activities were already in place? Did they have to be strengthened or re-organised?
67. Which MAMI activities had to be newly added?
68. Is active screening working well in the community? What screening criteria do you use?
69. How are community health workers/volunteers linking to the health facility? Explain.
70. How did community health workers perceive the extra tasks they were asked to do? Did they express concerns, and if so, what were they?

##### 4.3 Organisation of care in the health facility (evidence-based, continuity (referral), coordinated, integrated, comprehensive, people-centred, equipped, equity)

71. What activities are provided at the health facility, how, where by whom? Use the example table to answer.

Example table: Who delivers where what services in the primary healthcare centre

| Activities | How | Where | By whom |
|---|---|---|---|
| Sensitization on risks | | | |
| Health and nutrition promotion | | | |
| Screening (rapid assessment) | | | |
| IMNCI assessment, triage | | | |
| Anthropometry assessment | | | |
| MAMI risk assessment | | | |
| Feeding assessment | | | |
| Mental health assessment | | | |
| Classification and referral | | | |
| Treatment and support plan | | | |
| Enrolment | | | |
| Treatment and support | | | |
| Targeted counselling on feeding issues | | | |
| Targeted counselling on mental health issues | | | |
| Targeted counselling other (specify) | | | |
| Frequency of attendance decision for follow-up | | | |
| Referral in case of deterioration during enrolment | | | |
| Evaluate progress | | | |
| Evaluate outcome | | | |
| Referral in case of non-recovery at 6m | | | |
| Follow-up after exit | | | |

72. Which MAMI activities were already in place? Did they have to be strengthened or re-organised?
73. Which MAMI activities had to be newly added?
74. Is routine screening done in all health services and units frequented by infant-mother pairs? What screening criteria are used?
75. Was referral for maternal mental health possible?
76. How is referral to inpatient care organised for pairs whose status deteriorates, does it work well, or not?
77. How is counter-referral to outpatient care organised for pairs discharged from hospital, does it work well, or not?
78. What further support was most needed at 6 months?
79. Is there a follow up period after pairs exit at infant age 6m? If yes, for how long? and how is it organised?
80. Describe how are pairs are followed across services and in time (continuity of care).

##### 4.4 Organisation of staff: numbers, skill sets, sharing of tasks, supportive supervision, mentoring, job aids

81. Were sufficient number of skilled workers available to absorb MAMI? Explain.
82. What guidance or job aids did you use or develop? Explain.

83. Did you use v3 materials (if any) for organising and supporting health facility y implementation (job aids), and how?
84. How are clinical health workers linking, collaborating, sharing tasks, communicating on MAMI care at the health facility? Explain.
85. How are clinical health workers linking, communicating on MAMI care to other health facilities? Explain.
86. How organised and ready for quality implementation were you at the start (your opinion)? What went well, what went less well? Were roles and responsibilities clear for all implementers prior to starting? Explain.
87. Is supportive supervision and mentoring being provided? If yes, how is it organised, which tools are used?
88. How did health workers perceive to adopt the innovation/increase consistency/merge with what they were already doing? Specify for the different activities at the different levels.
89. How did clinical health workers perceive the extra tasks they were asked to do? Did they express concerns, and if so, what were they?

##### 4.5 Participation

90. Do you involve caregivers (community members) in care? Explain.
91. Prior to assessing risks and enrolling, did you ask the caregiver's perceived need and interest in receiving this service?
92. Were caregivers well informed and had a choice, were encouraged to take active part in care, how?
93. How did caregivers perceive the effort to return for follow-on visits? How do you motivate them?
94. Prior to assessing MAMI risks and enrolling pairs, did you ask the caregiver's perceived need and interest in receiving this service?
95. Did you assess the caregiver's satisfaction during and when exiting the MAMI Care Pathway?

##### 4.6 Partnerships

96. What is the role of the local health management system; how are MOH focal points involved in planning, supervising and improving quality, mentoring, evaluating?
97. Are there other technical partners providing support at the MAMI Sites? Who are they, what do they cover, how you collaborate?
98. Are there other technical partners providing support at the MAMI Sites? Who are they, what do they cover, how you collaborate?
99. Is there a communication or coordination system linking the various partners?

#### 5. *Monitoring and collaborative learning*

##### 5.1 Monitoring and reporting

100. Have you a monitoring system in place? If yes, to what degree you use existing data and systems?
101. List the indicators you report on monthly and give results for the period of reporting. Use the example table to answer.

Example table: Key indicators (country or site, period of reporting)

| | Total |
|---|---|
| **Sensitization** | |
| MAMI sensitization in the community (# of people reached) | |
| MAMI sensitization in the health facility (# of people reached) | |
| **Screening (rapid assessment)** | |
| Total pairs screened in the community | |
| Pairs screened at risk, referred for in-depth assessment | |
| Total pairs screened in the primary care facility | |
| Pairs screened at risk, referred for in-depth assessment | |
| **In-depth assessment** | |
| Total pairs assessed | |

| |
|---|
| a. Pairs assessed - male infant |
| b. Pairs assessed - female infant |
| Pairs assessed classified at moderate risk (yellow) |
| Pairs assessed classified at high risk (red) and referred |
| **Enrolment in outpatient care** |
| Total pairs newly enrolled |
| a. Pairs newly enrolled - male infant |
| b. Pairs newly enrolled - female infant |
| **Referral during outpatient care** |
| Total pairs referred to hospital |
| a. Pairs referred to hospital - infant high risk |
| b. Pairs referred to hospital - mother high risk |
| **Outcome of outpatient care** |
| Total pairs exited from the outpatient Care Pathway |
| Total pairs exited at infant age 6m |
| Pairs not recovered at infant age 6m and referred to continue care |
| a. Pairs not recovered at infant age 6m - infant special care |
| b. Pairs not recovered at infant age 6m - mother special care |
| Pairs recovered at infant age 6m |
| Total pairs exited before infant age 6m |
| Pairs died before the age of 6m |
| Pairs lost to follow up (defaulted) before the age of 6m |

Example table: MAMI enrolment by age group (country or site, period of reporting)

| | **Total** |
|---|---|
| **Age of infants at enrolment in outpatient care** | |
| <1 month | |
| 1–<2 months | |
| 2–<3 months | |
| 3–<4 months | |
| 4–<5 months | |
| 5–<6 months | |

102. Do you consolidate monthly monitoring data on service performance? Do you use digitized tools? Explain.

103. Do you consolidate individual data on assessment and enrolment? Do you use digitized tools? Explain.

104. Describe if and what qualitative data you collect, for what purpose, how you collect it, with what tools, and how you consolidate and report on them?

105. Do you capture lessons? Explain.

106. What key lessons have you learned that you think would be helpful for managing small and nutritionally at-risk infants u6m and their mothers?

107. What key successes you want to share?

108. What key challenges did you face? Which actions you have undertaken to overcome these, and did you succeed to overcome these, or not?

##### 5.2 Improving quality

109. Are monitoring results (data tables and figures and lessons) used for quality improvement (QI) to identify weaknesses in data collection and quality of care that needs improvement (e.g., in monthly meetings)? Explain.

110. Do you use adaptive management for quality improvement and learning (e.g., using the plan-do-verify-adapt cycle)? Explain.

111. What has MAMI added to your work and experience?

##### 5.3 Disseminating information and learning

112. How is in-country sharing of information on MAMI organized? Explain the different pathways.

113. How is wider sharing of information on MAMI organized, outside of the country? Explain the different pathways.

114. What learning methods or communication platforms are being used by your managers, by the implementers, and how did they come about? Explain.

115. Have you established a national learning and information sharing entity (e.g., community of practice, Country Chapter)? Explain.

116. Have you involved national research institutions in MAMI? Explain.

117. How did you explore their potential involvement in documenting lessons, evaluating evidence gaps and proposing research studies (including donors).

118. Is any evaluation in progress or planned? Explain.

119. Have you identified any research gaps? If so, what are they?

**5.4 Maintaining and sustaining quality services**

120. Are the MAMI activities that you implement sustainable? Explain.

121. How can the specific MAMI activities be made more sustainable? what are barriers and facilitators? Explain.

122. Are they resilient to shocks? Explain.

123. Can the specific MAMI activities be made more resilient? what are barriers and facilitators? Explain.

**5.5 Ensuring accountability to…**

124. Who are you accountable to, how and for what?

**5.6 Advocating for … strengthening services and adapting policies**

125. Are you engaging decision-makers, champions, gate-keepers in MAMI?

126. What advocating tools you use or have you developed to highlight the burden, the importance of addressing MAMI, the effectiveness of MAMI?

127. Are you involved/do you plan to engage in national policies, guidelines, strategies, processes for contributing to evidence and learning? If yes, in what way?

128. Is the accountability of MAMI in your implementation design sufficient, or what is missing, what should be strengthened and how?

#### *6. Recommendations*

129. List or describe changes you suggest for simplifying or improving the v3 materials.

130. List or describe additional resources you wish to have to improve planning, organizing, implementing, monitoring, learning, or expanding the evidence base.

131. What do you identify as most important gap / need that should be addressed, by whom and at what level?

132. Share any other general or specific recommendations you have?
