## Supplemental Material 2 Data Tools Planning for Scale-Up for "Reflective Learning from Implementing Care Pathways for Vulnerable Infants and Their Mothers: Case studies from Pakistan, South Sudan and Yemen"

### Data Tool – Planning for Successful Scale-Up of the MAMI Care Pathway Approach

| Questions related to potential scalability | Yes (+) | No (–) | More information / action needed |
|---|---|---|---|
| 1. Is input about the project being sought from a range of stakeholders (e.g. policy-makers, programme managers, providers, NGOs, beneficiaries)? | | | |
| Are individuals from the future implementing agency involved in the design and implementation of the pilot? | | | |
| Does the project have mechanisms for building ownership in the future implementing organization? | | | |
| 2. Does the innovation address a persistent health or service- delivery problem? | | | |
| Is the innovation based on sound evidence and preferable to alternative approaches? | | | |
| Given the financial and human-resource requirements, is the innovation feasible in the local settings where it is to be implemented? | | | |
| Is the innovation consistent with existing national health policies, plans and priorities? | | | |
| 3. Is the project being designed in light of agreed-upon stakeholder expectations for where and to what extent interventions are to be scaled-up? | | | |
| 4. Has the project identified and taken into consideration community, cultural and gender factors that might constrain or support implementation of the innovation? | | | |
| Have the norms, values and operational culture of the implementing agency been taken into account in the design of the project? | | | |
| Have the opportunities and constraints of the political, policy, health-sector and other institutional factors been considered in designing the project? | | | |
| 5. Has the package of interventions been kept as simple as possible without jeopardizing outcomes? | | | |
| 6. Is the innovation being tested in the variety of sociocultural and geographic settings where it will be scaled-up? | | | |
| Is the innovation being tested in the type of service-delivery points and institutional settings in which it will be scaled-up? | | | |
| 7. Does the innovation being tested require human and financial resources that can reasonably be expected to be available during scale-up? | | | |
| Will the financing of the innovation be sustainable? | | | |
| Does the health system currently have the capacity to implement the innovation? If not, are there plans to test ways to increase health-systems capacity? | | | |
| 8. Are appropriate steps being taken to assess and document health outcomes as well as the process of implementation? | | | |
| 9. Is there provision for early and continuous engagement with donors and technical partners to build a broad base of financial support for scale-up? | | | |
| 10. Are there plans to advocate for changes in policies, regulations and other health-systems components needed to institutionalize the innovation? | | | |
| 11. Does the project design include mechanisms to review progress and incorporate new learning into the implementation process? | | | |
| Is there a plan to share findings and insights from the pilot project during implementation? | | | |
| 12. Is there a shared understanding among key stakeholders about the importance of having adequate evidence related to the feasibility and outcomes of the innovation prior to scaling up? | | | |

WHO ExpandNet, 2011, Beginning with the end in mind: planning pilot projects and other programmatic research for successful scaling up.

1. Engage in a participatory process involving key stakeholders
2. Ensure the relevance of the proposed innovation
3. Reach consensus on expectations for scale-up
4. Tailor the innovation to the sociocultural and institutional settings
5. Keep the innovation as simple as possible
6. Test the innovation in the variety of sociocultural and institutional settings where it will be scaled-up
7. Test the innovation under the routine operating conditions and existing resource constraints of the health system
8. Develop plans to assess and document the process of implementation
9. Advocate with donors and other sources of funding for financial support beyond the pilot stage
10. Prepare to advocate for necessary changes in policies, regulations and other health-systems components
11. Develop plans for how to promote learning and disseminate information
12. Plan on being cautious about initiating scale-up before the required evidence is available
