## Supplemental Material 2 Data Tools Normalization for "Reflective Learning from Implementing Care Pathways for Vulnerable Infants and Their Mothers: Case studies from Pakistan, South Sudan and Yemen"

### Data Tool – Adopting the MAMI Care Pathway Approach (Normalization)

***Name of the responder and position:*** ____________________

***Date of response:*** ____________________

**Agency:** ____________________

**QUESTIONS Clinical Healthcare Worker – Key informant interview**

#### PRE-QUESTIONS

1. Please confirm, your name is […], your current position is […]
2. Where are you working, in which establishment, health facility?
3. Since how long do you work there? Give start date.
4. When was the MAMI Care Pathway introduced at your health facility? Give start date.
5. What is your function in relation to the MAMI Care Pathway?
6. (If started working after MAMI was introduced) Were you exposed to MAMI before joining the health facility, where, in what function?
7. (If started working after MAMI was introduced) Did you have specific MAMI knowledge and skills prior to joining the current position?

#### QUESTIONS

*Questions seek the opinion of the clinical health worker about implementing the MAMI Care Pathway in his/her setting versus to what they did before for small vulnerable infants and their mothers. Ask the respondent to explain their answer (f yes, explain how, if no, explain why not?) and give a grade on a Likert scale from 0 (not at all) to 5 (completely):*

##### Coherence–meaning and sense making

1. Is the MAMI Care Pathway easy to describe? Can you appreciate how it differs from current ways of working, from what you did before to support small vulnerable infants and their mothers?
   **Participants distinguish the intervention from current ways of working:** not at all to completely

2. Have you and your colleagues a common understanding of the aims, objectives and expected outcomes of the MAMI Care Pathway?
   **Participants collectively agree about the purpose of the intervention:** not at all to completely

3. Do you understand what implementing the MAMI Care Pathway requires from you (specific tasks and responsibilities)?
   **Participants individually understand what the intervention requires of them:** not at all to completely

4. Can you easily grasp the potential value, benefits and importance of the MAMI Care Pathway?
   **Participants construct potential value of the intervention for their work:** not at all to completely

##### Cognitive participation–commitment and engagement

5. Are you (or other key individual) able and willing to get others involved in the MAMI Care Pathway? Are you actively engaged in making the MAMI Care Pathway work in your setting?
   **Key individuals drive the intervention forward:** not at all to completely

6. Do you believe and agree that being involved is right, and that by accepting the MAMI Care Pathway as part of your work you contribute to its implementation?
   **Participants agree that the intervention should be part of their work:** not at all to completely

7. Do you have the capacity and are you willing to organise you and your colleagues and collectively contribute to the work involved for implementing the MAMI Care Pathway?
   **Participants buy in to the intervention:** not at all to completely

8. Do you have the capacity and are you willing to collectively define the actions and procedures needed to keep the practice ongoing (invest your time, energy to keep it going)?
   **Participants continue to support the intervention:** not at all to completely

##### Collective action–work done to enable the intervention to happen

9. Are you and colleagues able to do the tasks required to implement the MAMI Care Pathway (to operationalise its components in practice)?
   **Participants perform the tasks required by the intervention:** not at all to completely

10. Do you maintain trust in the intervention and in each other's work and expertise in implementing the MAMI Care Pathway?
    **Participants maintain their trust in the intervention and in each other:** not at all to completely

11. Is the work required for implementing the MAMI Care Pathway distributed to participants with the right mix of skills and training? Did it impact on the division of labour, resources, power, responsibilities between colleagues (tasks and skill sharing)? Was extensive training needed before implementing the MAMI Care Pathway? (originally Q13)
    **The work of the intervention is appropriately allocated to participants:** not at all to completely

12. Is the implementation of the MAMI Care Pathway adequately supported by the advisor/manager?
    **The intervention is adequately supported by its host organisation:** not at all to completely

##### Reflective monitoring–reflect on or appraise the benefits

13. Do you have access to information on the quality of care and outcome of the MAMI Care Pathway (monitoring and evaluation information)?
    **Participants access information about the effects of the intervention:** not at all to completely

14. Do you collectively agree on the quality of care and the effects of the MAMI Care Pathway because of formal monitoring?
    **Participants collectively assess the intervention as worthwhile:** not at all to completely

15. Do you individually think the MAMI Care Pathway is worthwhile?
    **Participants individually assess the intervention as worthwhile:** not at all to completely

16. Can you make changes to the intervention as individual or group in response to the appraisal?
    **Participants modify their work in response to their appraisal of the intervention:** not at all to completely
